# Affinity proteomics-based non-invasive detection of clinically significant liver disease

**DOI:** 10.1101/2025.06.13.25329564

**Authors:** Sriram Balasubramani, Anna Sophie Karl, Julia Alexandra Borchert, Christina Schrader, Malin Fromme, Can Kayatekin, Bailin Zhang, Mikhail Levit, Pavithra Krishnaswami, Louise E. van Eekeren, Leo A. B. Joosten, Petra Tomanová, Katharina Remih, Pavel Strnad

## Abstract

**Background:** Since liver disease is often clinically unapparent, non-invasive biomarkers predicting future major adverse liver outcomes (MALO) are urgently needed. Therefore, we assessed the usefulness of a novel, proximity extension assay (PEA)-based high-throughput targeted proteomics method to predict MALOs.

**Methods:** PEA plasma proteomic data (>2900 proteins) and clinical information were accessed from the population-based UK Biobank (UKB) cohort, including >53000 individuals with a median follow-up of >10 years and its subcohorts of obese (>12900) and diabetic (>1600) participants. The validation cohorts comprised 287 subjects with severe alpha1-antitrypsin deficiency (AATD), Pi*ZZ genotype, and 960 people living with HIV (PLHIV), who underwent liver stiffness measurement (LSM) via transient elastography. Selected PEA parameters were compared to routine measurements. Bayes-moderated linear models (age and sex as covariates) assessed the differential abundance. Logistic regression was used to identify and validate a novel prognostic score.

**Results:** Routine gamma-glutamyltransferase (GGT) and aspartate aminotransferase (AST) levels strongly correlated with PEA-based measurements (r=0.91 and r=0.68, respectively). Similarly, PEA-based thrombospondin-2 levels strongly correlated with immunoassay-based values (r=0.85). Twenty proteins were consistently associated with future MALOs/increased LSM in all cohorts. UKB cohort was used to develop a novel five-component PEA score that demonstrated superior predictive power (AUROC=0.84) compared to established indices/scores, including AST-to-platelet-ratio index (APRI, AUROC=0.73) and Fibrosis-4 index (FIB4, AUROC=0.72) and its attractive predictive power was sustained in diabetic/obese subcohorts. In PLHIV and AATD validation cohorts, all five components showed gradual increases across fibrosis stages, and PEA score numerically outperformed APRI/FIB4 in predicting significant liver disease.

**Conclusion:** Our study identifies a new PEA score consisting of epithelial/hepatic stellate cell markers that demonstrates an attractive discriminative ability in several independent cohorts and different liver disease aetiologies.

## Introduction

Although potentially preventable, liver-related mortality constitutes the second leading cause of lost working life in Europe[1]. To make things worse, liver disease is often asymptomatic and becomes only apparent when major adverse liver outcomes (MALO) such as ascites, variceal bleeding, or HCC occur. However, such a late diagnosis is associated with poor prognosis[1]. Therefore, screening for liver disease is of utmost importance, especially in subjects at increased risk, such as obese individuals or patients with diabetes[2]. Serum liver enzymes, *i.e.,* aspartate/alanine aminotransferase (AST/ALT) or gamma-glutamyltransferase (GGT), are commonly used; however, they reflect a short-term damage rather than the long-term remodelling known as fibrosis that is of key prognostic relevance[1,3]. For the latter, several serum marker-based scores, such as Fibrosis-4 (FIB4), have been developed that often combine liver injury markers with platelet count as a marker of advanced liver fibrosis with portal hypertension[4]. Alternatively, liver stiffness measurement (LSM) via ultrasound or magnetic resonance imaging is used as a surrogate of liver scarring[5]. Since the liver represents a key secretory organ responsible for producing most of the proteins found in the bloodstream, serum/plasma proteomics constitutes an attractive and emerging approach to identifying new biomarkers[6,7]. In addition to the traditional, mass spectrometry-based approach, several commercially available, affinity-based platforms have been developed and offer both high-throughput as well as worldwide standardisation[8,9]. Among them, the aptamer-based SomaScan technology and the proximity extension assay (PEA)-based Olink® platform are the most widely used[10,11]. The latter has been extensively studied in the UK Biobank (UKB), a large-scale community-based, prospective cohort, where it yielded multiple valuable insights into human health[12,13]. In contrast, the applicability of this technique in hepatology remains less well explored. To change that, we assessed the usefulness of the PEA technology to predict MALOs in the UKB cohort. We assessed the whole proteomic UKB cohort as well as subcohorts of participants with obesity and type-2 diabetes (DM2), as established high-risk groups that may particularly benefit from liver disease screening. The findings were validated in cohorts of patients with severe alpha1-antitrypsin deficiency (AATD), an established proteotoxic liver disease[14,15] and people living with HIV (PLHIV)[16,17]. The performance of the novel PEA score was compared to values obtained with alternative techniques, including the ©LiverRisk score that was validated in multiple independent cohorts[18].

## Materials and Methods

### Patient cohorts

#### UKB Cohort

The UKB comprises a large-scale, community-based prospective study that recruited ∼500,000 participants with a mean follow-up period of 13.5 years[19]. The baseline dataset includes demographic data (age, sex, ethnicity), clinical information (BMI, comorbidities such as DM2) and laboratory values. Baseline visits took place between 2006-2010 in 22 centres across the UK. Inpatient hospital records starting in 1996 were used to obtain diagnoses based on the International Classification of Diseases version 10 (ICD-10) and operations/procedures performed based on operations/procedure codes version 4 (OPCS-4). Mortality data, including date/cause of death, were accessed through national death registries to analyse the associations between proteomic biomarkers and overall/liver-related (primary cause of death stated with ICD-10 codes K70-K77) mortality. All data handling and analyses were performed under the guidelines and regulations approved by the UKB, under application number 148742.

#### AATD cohort

The cohort consisted of 287 plasma samples collected from individuals with genetically confirmed homozygous Pi*Z mutation in alpha1-antitrypsin gene (Pi*ZZ) as part of the European Alpha1 liver study[20–22]. All participants underwent thorough liver/lung phenotyping by a standardised work-up comprising questionnaires and laboratory analyses, including standard serological testing with liver enzymes or platelet count. LSM via transient elastography [TE, (FibroScan®, Echosens, Paris, France)] was performed by experienced medical staff. The examinations were conducted between 2015-2023. Inclusion criteria for participation were (i)adolescence (age ≥18 years), (ii)no pregnancy, and (iii)ability to provide written informed consent. Exclusion criteria were previous liver transplantation and inability to meet criteria described above. Plasma thrombospondin-2 (THBS2) levels were quantified with an ELISA kit (DTSP20, R&D Systems, Abingdon, UK). The Ethics Committee of Aachen University (Aachen: EK 173/15) provided ethical approval, and all participants gave written informed consent. Everyone was assessed following the ethical guidelines of the Helsinki Declaration (Hong Kong amendment) and Good Clinical Practice (European guidelines).

#### PLHIV cohort

The previously published PLHIV cohort consists of 960 participants of the 2000HIV study[16,17]. At the baseline visit, they underwent a standardised examination comprising questionnaires, blood sampling, LSM via TE, and clinical data extraction from medical records. Laboratory parameters used for clinical assessments were obtained from available clinical data.

### Definition of patient subgroups

Patients with MALOs were identified from UKB using a combination of ICD-10/OPCS-4 codes as well as cancer/death registries. We used ICD-10 codes suggesting the presence of liver cirrhosis, HCC, chronic liver failure, portal hypertension and oesophageal varices. OPCS-4 codes referring to management of variceal bleeding, portal hypertension and ascites due to hepatic causes were also taken into account. The criteria are based on the selection of cases by Innes *et al.*[23] with minor modifications (Table S1). 80 patients who received a liver transplantation before their baseline assessment were excluded.

In both the AATD and PLHIV cohorts, LSMs were used as a surrogate for liver fibrosis and to classify patients according to established cut-offs. Participants were categorised as having no/minimal fibrosis (LSM <7.0 kPa), significant fibrosis (LSM ≥7.0 kPa), advanced fibrosis (LSM ≥10.0 kPa), or cirrhosis with potential portal hypertension (LSM >15.0 kPa) based on previously validated thresholds[20–22,24].

### Plasma proteomic data

Plasma samples were collected from participants in all three cohorts and assessed using PEA technology. All proteomic data are reported as dimensionless normalised protein expression (NPX) values[12].

For the UKB cohort, samples from 54219 participants who were part of the UKB Pharma Proteomics Project, including 46595 randomly selected UKB participants from baseline assessments, 1268 participants belonging to the COVID-19 repeat image study (with multiple visits), and 6376 participants directly selected by consortium members, were assessed with the Olink® Explore platform, capturing 2,923 unique proteins. After excluding 80 patients with previous liver transplantation, proteomic data from 52,998 participants were included in the analysis.

For the AATD cohort, plasma samples from 287 subjects with Pi*ZZ genotype were analysed using the Olink® Explore HT platform, which provided coverage of 5,416 unique proteins. The PLHIV cohort contributed plasma samples from 960 subjects that were analysed using the Olink® Explore platform, capturing 2,923 unique proteins, as described [16].

For further methods, please refer to supplement.

## Results

Our study aimed to test the usefulness of a PEA proteomics to predict significant liver disease in three independent cohorts: i) the community-based UKB cohort; ii) a multi-centre AATD cohort; and iii) a PLHIV cohort (Fig. 1A). 52998 participants from UKB with available routine lab and PEA measurement, representative of the entire UKB cohort, were assessed (Table 1). Two liver-related biomarkers (GGT/GGT1 and AST/GOT1) were quantified by both routine methods and PEA and their measurements displayed a strong correlation (r=0.68 and r=0.91, respectively, Fig. 1B). Among the participants with available PEA data, 495 developed a MALO (Fig. 1A, Table 1). Given that the occurrence of MALOs in this community-based cohort was low (*i.e.,* <1%), we studied the performance of PEA proteomics in two higher-risk subcohorts, *i.e.,* patients with baseline obesity (BMI ≥30) and DM2. The subcohorts encompassed 12965 (obesity) and 1608 (DM2) subjects who developed 228 (1.8%) and 73 (4.5%) MALOs, respectively (Fig. 1A, Tables S2-S3).

**Fig. 1.**
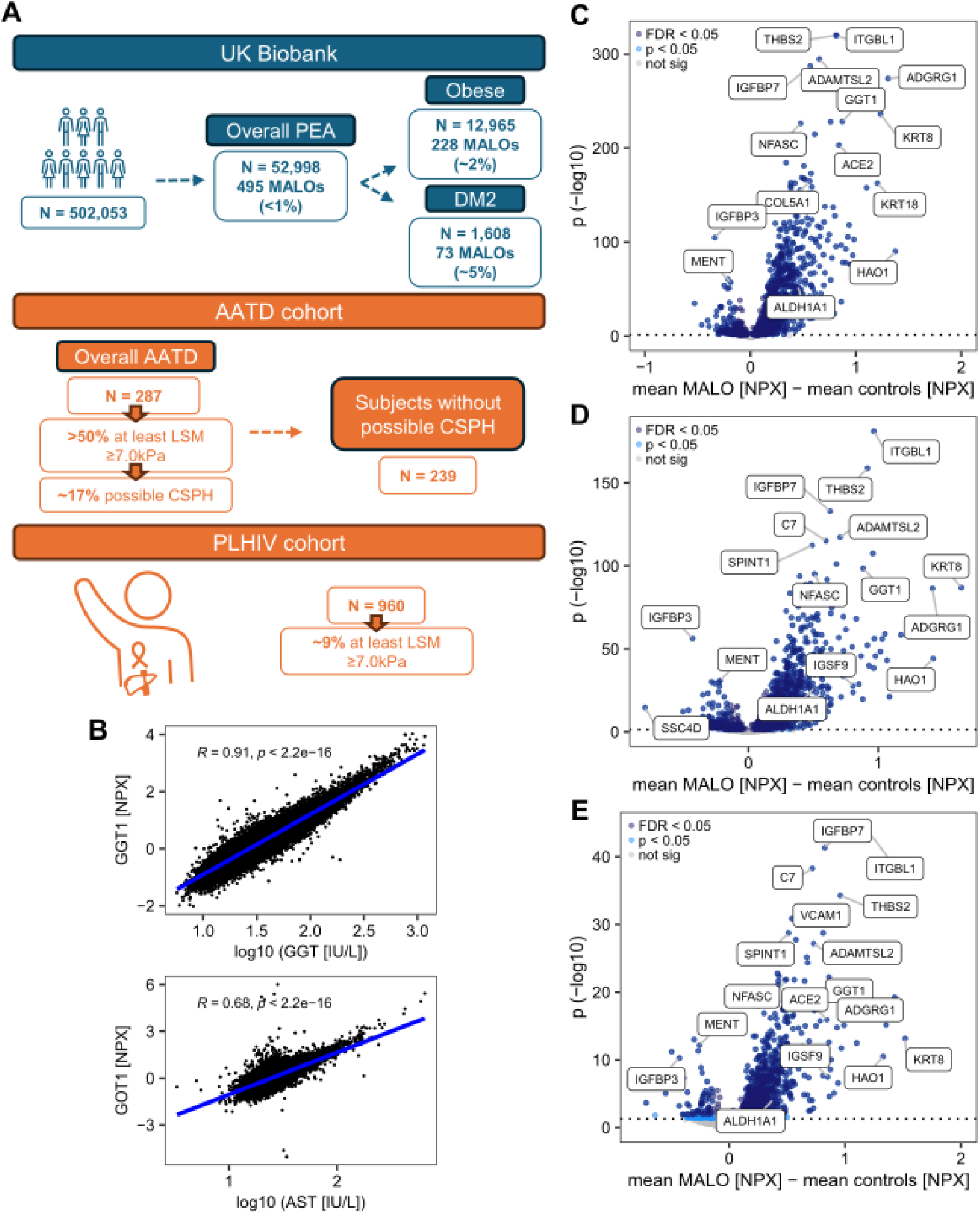
Proximity extension assay (PEA)-based proteomics in participants from the UK Biobank cohort (UKB) with/without major adverse liver outcomes (MALOs). A: Overview of the analysed cohorts. B: Correlation between routine and PEA-based measurements of gamma-glutamyltransferase (PEA: GGT1; routine: GGT) and aspartate aminotransferase (PEA: GOT1, routine: AST). Spearman rank correlation coefficients and corresponding *p* values are shown. C-E: Volcano plots of differentially abundant proteins between subjects with/without future MALOs in the entire UKB cohort with available PEA analysis [C], in a subgroup of obese (BMI ≥30) participants [D] and a subgroup with type-2 diabetes (DM2) [E]. The dotted horizontal line reflects a *p* =0.05. *AATD: alpha1-antitrypsin deficiency; possible CSPH: clinically significant portal hypertension, refers to LSM >15.0 kPa; GOT1: glutamic-oxaloacetic transaminase 1; LSM: liver stiffness measurement; PLHIV: people living with HIV*.

**Table 1:** Demographic and routine parameters of UK Biobank (UKB) cohorts. The entire UKB cohort and its subset with available proteomic data (PEA cohort) are shown, the latter being subdivided into subjects who did vs. did not develop major adverse liver outcomes during the follow-up (MALO/non-MALO). Data are expressed as median (25^th^-75^th^ percentile) for continuous variables and n (%) for categorical variables. *p* values are calculated with ANOVA (*without covariates, **with covariates age, sex, and BMI). Parameters with very low *p* values are shown as <1.91E-254. Parameters with *p* <1.00E-100 are highlighted in bold. *ALT: alanine aminotransferase; ALP: alkaline phosphatase; AST: aspartate aminotransferase; GGT: gamma-glutamyltransferase; HbA1C: haemoglobin A1c; PEA: proximity extension assay*.

| Characteristics | UKB cohort,<br>N=502,053 | PEA cohort,<br>N=52,998 | PEA cohort |  | *p* value<br>(A vs B) |
| --- | --- | --- | --- | --- | --- |
|  |  |  | Non-MALOs (A)<br>N=52,503 | MALOs (B)<br>N=495 |  |
| Age (years) | 58 (50-63) | 58 (50-64) | 58 (50-64) | 61 (56-65) | 3.71E-17* |
| Sex (Female) | 273,132 (54%) | 28,571 (54%) | 28,394 (54%) | 177 (36%) | 3.88E-16* |
| BMI (kg/m<sup>2</sup>) | 26.7 (24.1-29.9) | 26.8 (24.2-29.9) | 26.8 (24.2-29.9) | 29.2 (25.8-33.1) | 1.04E-31* |
| Type-2 Diabetes | 13,656 (2.7%) | 1,608 (3.0%) | 1,535 (2.9%) | 73 (15%) | 3.42E-52** |
| Blood glucose<br>(mmol/L) | 4.93 (4.60-5.32) | 4.94 (4.60-5.33) | 4.93 (4.60-5.33) | 5.18 (4.67-5.89) | 1.60E-33** |
| HbA1C<br>(mmol/mol) | 35.2 (32.8-37.9) | 35.3 (32.9-38.1) | 35.3 (32.9-38.1) | 36.9 (33.5-41.4) | 1.54E-26** |
| Total protein (g/L) | 72.3 (69.7-75.1) | 72.3 (69.7-75.1) | 72.3 (69.7-75.0) | 73.1 (70.1-76.9) | 6.39E-08** |
| Albumin (g/L) | 45.20 (43.49-46.92) | 45.13 (43.40-46.88) | 45.14 (43.41-46.89) | 43.74 (41.73-45.93) | 1.20E-36** |
| **ALT (IU/L)** | 20 (15-27) | 20 (15-27) | **20 (15-27)** | **26 (18-44)** | **3.05E-101\*\*** |
| **AST (IU/L)** | 24 (21-29) | 24 (21-29) | **24 (21-29)** | **32 (24-46)** | **1.91E-254\*\*** |
| **GGT (IU/L)** | 26 (19-41) | 26 (19-41) | **26 (19-41)** | **66 (32-159)** | **<1.91E-254\*\*** |
| ALP (IU/L) | 80 (67-96) | 81 (67-96) | 81 (67-96) | 94 (77-117) | 1.72E-95** |
| Total bilirubin<br>(μmol/L) | 8.1 (6.4-10.4) | 8.1 (6.4-10.4) | 8.1 (6.4-10.4) | 9.4 (6.9-12.7) | 1.51E-25** |
| **Direct bilirubin**<br>**(μmol/L)** | 1.61 (1.30-2.09) | 1.62 (1.30-2.11) | **1.62 (1.30-2.10)** | **2.09 (1.52-3.05)** | **8.59E-116\*\*** |
| Platelets (10<sup>9</sup>/L) | 248 (213-287) | 248 (213-287) | 248 (213-287) | 220 (172-265) | 1.58E-28** |

In all assessed cohorts, participants developing MALOs were older, more often male, and displayed higher levels of routinely assessed liver function tests (Tables 1, S2-S3). After correction for multiple testing (false discovery rate (FDR)-adjusted *p* value <0.05), differential abundance analysis (age and sex as covariates) revealed 1772 differentially abundant proteins in the overall cohort, and 1704 and 815 proteins in the obese and diabetic subcohorts (Fig. 1C-E; Tables S4-S6).

### Proteomic signatures associated with advanced liver disease across UKB cohorts

The overall pattern was similar in all assessed cohorts, with most proteins showing higher levels in subjects with vs. without future MALOs (Fig. 1C-E). Only 15.1%, 19.0% and 2.4% of significantly altered proteins were diminished in the overall, obese and diabetic subcohorts with future MALOs (Fig. 1C-E). 796 proteins were significantly altered in all analyses (Fig. 2A). When the top 100 differentially abundant proteins were assigned to their tissue of origin, proteins produced by the liver, intestine, or lymphoid tissues were the most common hits, while approximately 25% displayed low tissue specificity (Fig. S1).

**Fig. 2.**
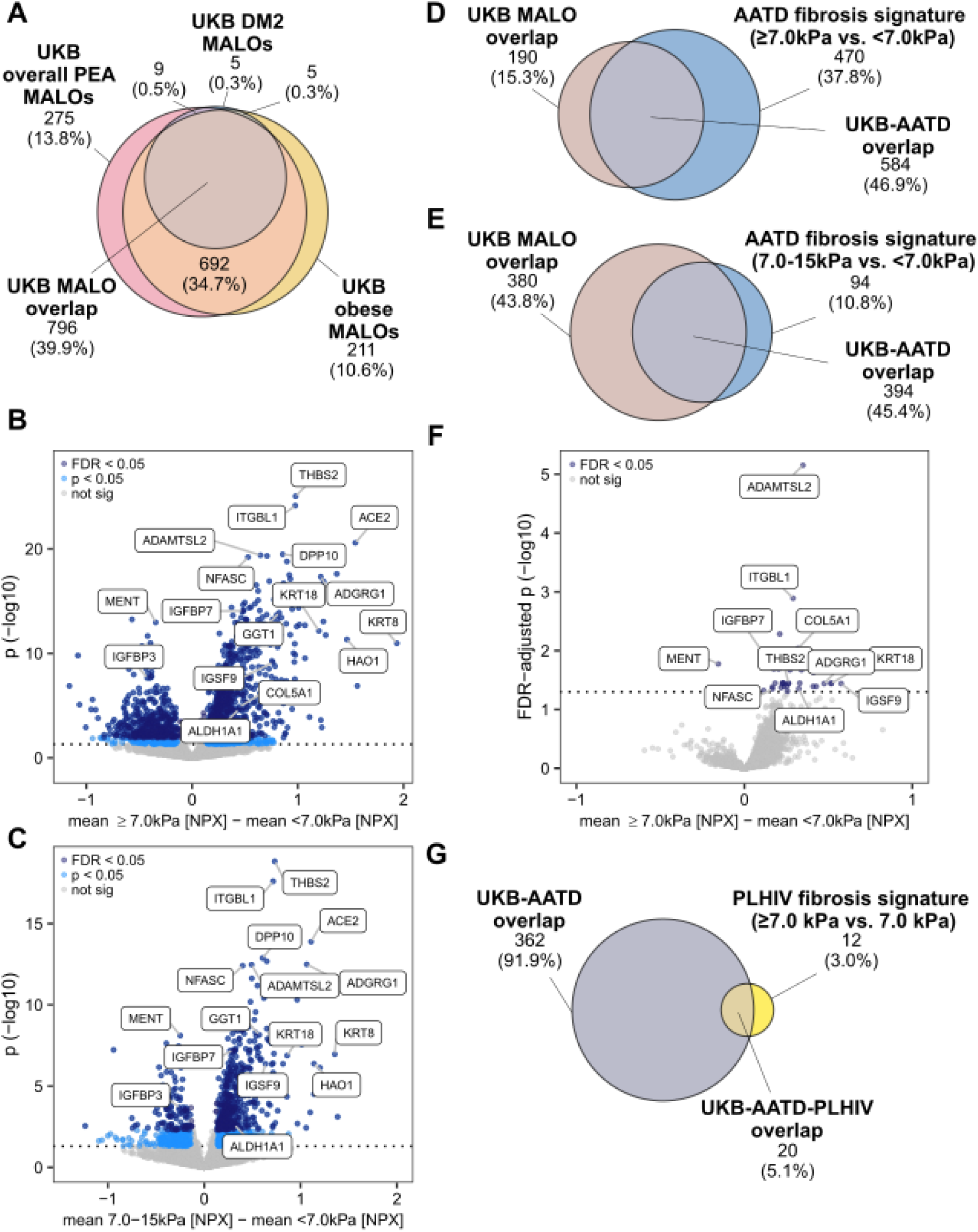
Identification of proteins consistently associated with significant liver disease across studied cohorts. A: Venn diagram displays the overlap between signatures of proteins associated with MALO in three different subcohorts of the UKB (overall PEA cohort and subgroups of obese patients or patients with type-2 diabetes (DM2)). 796 proteins were altered (false discovery rate (FDR) <0.05) in all subcohorts. B/C: Volcano plots displaying the results of the differential abundance analyses between patients with alpha1-antitrypsin deficiency (AATD) with significant liver fibrosis (B: LSM ≥7.0 kPa, C: LSM between 7.0-15.0 kPa, *i.e.,* excluding patients with possible clinically significant portal hypertension) and those without (LSM <7.0 kPa). 1266 and 548 proteins were differentially abundant (FDR <0.05), respectively. D/E: Overlaps between the proteomic signatures presented in B and C and the MALO-associated features identified in the UKB shown as Venn diagrams. F: Volcano plot visualises differentially abundant plasma proteins in people living with HIV (PLHIV) with high (≥7.0 kPa) vs. low (<7.0 kPa) LSM values as surrogates of significant liver fibrosis. 32 proteins were differentially abundant (FDR <0.05). G: Overlaps between proteomic signatures detected in the PLHIV cohort and the proteins that significantly differed in all previous analyses (UKB-AATD overlap) are presented in a Venn diagram. 20 proteins were differentially abundant in all analyses. *LSM: liver stiffness measurement; MALO: major adverse liver outcomes; PEA: proximity extension assay; UKB: the UK Biobank*.

### Validation of proteomic signatures in AATD and PLHIV cohorts

To discover aetiology-independent biomarkers, we turned to a cohort of 287 subjects with severe AATD (Pi*ZZ genotype [14,15]), whose liver fibrosis stage was characterised via LSM as well as clinically (Table 2). Although the baseline demographic parameters were comparable, subcohorts with higher fibrosis stages were more often male, had higher BMI, elevated liver enzymes and lower platelets (Table 2). Similar to UKB, PEA-based GGT1/GOT1 strongly correlated with routine GGT/AST measurements (r=0.90 and r=0.87, Fig. S2A-B). A comparison of subjects with vs. without any clinically significant fibrosis (LSM ≥7.0 kPa vs. <7.0 kPa) yielded 1266 significantly altered proteins (FDR <0.05) (Fig. 2B; Table S7). To study less advanced disease, we analysed the AATD subcohort without portal hypertension, *i.e.,* LSM ≤15.0 kPa. This assessment yielded 548 significantly altered (FDR <0.05) proteins (Fig. 2C; Table S8). The biomarkers identified in the AATD cohort strongly overlapped with the proteins found in the UKB analyses (*i.e.,* 584 and 394 overlapping proteins (Fig. 2D-E)). Unlike the AATD cohort, the PLHIV cohort had only a limited number of subjects with significant fibrosis (LSM ≥7.0 kPa; n=86; 9.0%; Table S9), and only 32 proteins differed significantly (FDR <0.05) between the subgroups with high vs. low LSM (Fig. 2F). 20 of them, *i.e.,* the majority, were also significantly altered in all previous analyses. (Fig. 2G). Out of them, 19 were constantly increased and one was constantly decreased (MENT: C1orf56, Chromosome 1 open reading frame 56). Mapping these proteins to publicly available liver single-cell sequencing data (Table 3) revealed that many are related to hepatic stellate cells (HSC) or epithelial cells (likely reflecting hepatocellular injury). Not surprisingly, the correlations between the biomarkers were weakest when the entire UKB cohort was considered and increased in cohorts enriched for subjects with liver disease (Figs. S3-S7). As expected, MENT displayed negative correlations while other biomarkers correlated positively (Figs. S3-S7). Hepatocyte and HSC-related biomarkers displayed stronger correlations with each other, while weaker correlations were seen between markers from different cellular sources. When assessing the association with overall/liver-related death, all biomarkers but MENT presented with elevated odds ratios. Although most of them were significantly associated with overall death, the association with liver-related death was constantly stronger (Figs. S8-S9). To further validate the reliability of PEA quantification, we assessed THBS2 levels in the AATD cohort with a commercially available ELISA. Both measurements displayed a strong correlation (r=0.88, Fig. S2C), and the PEA assessment yielded a somewhat higher area under the receiver operating curve (AUROC) for discriminating subjects with vs. without significant fibrosis (*i.e.,* LSM ≥7.0 kPa vs. <7.0 kPa; Fig. S2D).

**Table 2:** Demographic and routine parameters of subjects with severe alpha1-antitrypsin deficiency (Pi*ZZ genotype). Categorization of patients is based on liver stiffness measurement (LSM), expressed in kPa. The subgroup ≥25 includes 19 subjects with decompensated liver cirrhosis. Data are expressed as median (25^th^-75^th^ percentile) for continuous variables and n (%) for categorical variables. *p* values are calculated with ANOVA (*without covariates, **with covariates age, sex, and BMI). *AATD: alpha1-antitrypsin deficiency; ALT: alanine aminotransferase; ALP: alkaline phosphatase; AST: aspartate aminotransferase; GGT: gamma-glutamyltransferase; Pi*ZZ: homozygous Pi*Z mutation in alpha1-antitrypsin gene*.

| Characteristics | N | AATD Pi*ZZ subjects, LSM (kPa) |  |  |  |  | *p* value |
| --- | --- | --- | --- | --- | --- | --- | --- |
| | | <7.0,<br>N=135 | 7.0-<10,<br>N=55 | 10-<15,<br>N=48 | 15-<25,<br>N=19 | $\geq 25$,<br>N=30 | |
| Age (years) | 285 | 56 (50-61) | 58 (51-65) | 59 (48-66) | 61 (55-63) | 60 (54-65) | 0.273* |
| Sex (Female) | 287 | 55 (41%) | 11 (20%) | 9 (19%) | 2 (11%) | 7 (23%) | 1.58E-03* |
| BMI (kg/m<sup>2</sup>) | 285 | 24.8 (22.9-27.0) | 27.3 (23.6-29.9) | 26.5 (24.6-29.3) | 25.6 (25.3-29.9) | 26.7 (24.3-30.5) | 0.019* |
| Diabetes (n) | 283 | 3 (2.2%) | 1 (1.9%) | 5 (11%) | 2 (11%) | 5 (17%) | 5.58E-03** |
| ALT (IU/L) | 284 | 31 (22-42) | 37 (29-63) | 43 (32-53) | 53 (41-62) | 57 (44-72) | 1.29E-04** |
| AST (IU/L) | 282 | 30 (23-36) | 35 (30-50) | 38 (32-45) | 49 (38-69) | 70 (49-96) | 3.92E-25** |
| GGT (IU/L) | 282 | 27 (21-42) | 54 (34-89) | 81 (42-129) | 94 (53-264) | 169 (87-301) | 4.92E-19** |
| ALP (IU/L) | 283 | 71 (60-85) | 87 (65-104) | 88 (72-102) | 94 (74-139) | 128 (104-189) | 9.12E-20** |
| Total bilirubin (μmol/L) | 280 | 8 (6-11) | 11 (9-13) | 11 (8-17) | 13 (9-19) | 22 (14-31) | 2.31E-11** |
| Platelets (10<sup>9</sup>/L) | 274 | 238 (196-278) | 204 (166-242) | 199 (134-242) | 185 (123-248) | 125 (101-150) | 5.59E-18** |

**Table 3:** Overview of proteins consistently associated with liver disease in all studied cohorts. ↑/↓ indicates proteins increased/decreased in diseased individuals. Significance levels are: ↑/↓ *p* <0.05; ↑↑/↓↓ *p* <0.001; ↑↑↑/↓↓↓ *p* <0.0001. Proteins are mapped to publicly available liver single-cell sequencing data[44]. *AATD: alpha1-antitrypsin deficiency; ACY1: aminoacylase 1; ADAMTSL2: ADAMTS-like protein 2; ADGRG1: adhesion G protein-coupled receptor G1; ALDH1A1: aldehyde dehydrogenase 1A1; ANGPT2: angiopoietin 2; CD80: CD80 molecule; CDH2: cadherin 2; CDH15: cadherin 15; CLSTN2: calsyntenin 2; DM2: type-2 diabetes; DSC2: desmocollin 2; ENG: endoglin; ENPP2: ectonucleotide pyrophosphatase/phosphodiesterase 2; HSC: hepatic stellate cell; IGFBP7: insulin-like growth factor-binding protein 7; IGSF9: Immunoglobulin superfamily member 9; ITGBL1: integrin beta-like protein 1; KRT18: keratin-18; LSM: liver stiffness measurement, expressed in kPa; MALO: major adverse liver outcomes; MENT: C1orf56 (chromosome 1 open reading frame 56); NA: not available; NFASC: neurofascin; NK: natural killer cell; PLHIV: people living with HIV; TFPI2: tissue factor pathway inhibitor 2; THBS2: thrombospondin-2; UKB: the UK Biobank cohort*.

| | MALO vs. non-MALO (UKB) | MALO vs. non-MALO (UKB obese) | MALO vs. non-MALO (UKB DM2) | LSM $\geq 7.0$ kPa vs. $< 7.0$ kPa (AATD) | LSM $\geq 7.0$ kPa vs. $< 7.0$ kPa (PLHIV) | Liver cell subtype |
| --- | --- | --- | --- | --- | --- | --- |
| THBS2 | ↑↑↑ | ↑↑↑ | ↑↑↑ | ↑↑↑ | ↑ | Activated HSC |
| ITGBL1 | ↑↑↑ | ↑↑↑ | ↑↑↑ | ↑↑↑ | ↑ | Activated HSC |
| ADAMTSL2 | ↑↑↑ | ↑↑↑ | ↑↑↑ | ↑↑↑ | ↑↑↑ | Activated HSC |
| IGFBP7 | ↑↑↑ | ↑↑↑ | ↑↑↑ | ↑↑↑ | ↑ | Activated HSC |
| ADGRG1 | ↑↑↑ | ↑↑↑ | ↑↑↑ | ↑↑↑ | ↑ | NK cell |
| NFASC | ↑↑↑ | ↑↑↑ | ↑↑↑ | ↑↑↑ | ↑ | HSC |
| KRT18 | ↑↑↑ | ↑↑↑ | ↑↑↑ | ↑↑↑ | ↑ | Epithelial |
| CD80 | ↑↑↑ | ↑↑↑ | ↑↑↑ | ↑↑↑ | ↑ | Kupffer cell |
| CLSTN2 | ↑↑↑ | ↑↑↑ | ↑↑↑ | ↑↑↑ | ↑ | Non-specific |
| ENPP2 | ↑↑↑ | ↑↑↑ | ↑↑↑ | ↑↑ | ↑ | Kupffer cell |
| CDH2 | ↑↑↑ | ↑↑↑ | ↑↑↑ | ↑↑↑ | ↑ | Epithelial |
| ACY1 | ↑↑↑ | ↑↑↑ | ↑↑↑ | ↑ | ↑ | Epithelial |
| ENG | ↑↑↑ | ↑↑↑ | ↑↑↑ | ↑ | ↑ | Endothelial |
| IGSF9 | ↑↑↑ | ↑↑↑ | ↑↑↑ | ↑↑↑ | ↑ | T cell |
| ANGPT2 | ↑↑↑ | ↑↑↑ | ↑↑ | ↑ | ↑ | Endothelial |
| DSC2 | ↑↑↑ | ↑↑↑ | ↑↑↑ | ↑↑↑ | ↑ | Mesothelial |
| MENT | ↓↓↓ | ↓↓↓ | ↓↓↓ | ↓↓↓ | ↓ | NA |
| TFPI2 | ↑↑↑ | ↑↑↑ | ↑↑↑ | ↑↑↑ | ↑ | Mesothelial |
| ALDH1A1 | ↑↑↑ | ↑↑↑ | ↑↑ | ↑ | ↑ | Epithelial |
| CDH15 | ↑↑↑ | ↑↑↑ | ↑ | ↑ | ↑ | Non-specific |

### Development and validation of a prognostic PEA model and its components

We then took the 20 consistently altered biomarkers and performed a multivariable logistic regression to obtain a score reliably predicting future MALOs in the entire UKB cohort (termed PEA score; Table S10; Figs. S10-S11, Table 4). For the model development, we systematically evaluated all possible combinations among the 20 biomarkers and decided to use a 5-feature signature as our analyses yielded an attractive predictive performance that was not substantially increased by the addition of further parameters (not shown). In the overall UKB cohort, the PEA score achieved an AUROC of 0.84 for predicting MALOs, which was superior to previously described indices/scores such as AST-to-platelet-ratio index (APRI, AUROC=0.73) or FIB4 (AUROC=0.72, Fig. 3A), and the same was true in the obese/diabetic UKB subcohorts (Fig. 3B-C). In the AATD and PLHIV validation cohorts, the PEA score maintained its superior performance compared to APRI/FIB4 and achieved similar or slightly better performance than the ©LiverRisk score (Fig. 3D-F). An assessment of the five components of the PEA score; integrin beta-like protein 1 (ITGBL1), ADAMTS-like protein 2 (ADAMTSL2), insulin-like growth factor-binding protein 7 (IGFBP7), keratin-18 (KRT18), and aldehyde dehydrogenase 1A1 (ALDH1A1), revealed stronger correlations between the HSC-related (*i.e.*, ITGBL1, ADAMTSL2, and IGFBP7) and epithelial markers (*i.e.,* ALDH1A1 and KRT18) but somewhat less pronounced associations between the groups (Figs. S3-S7). Notably, the levels of all biomarkers significantly differed among subjects with/without future MALOs as well as with lower vs. higher liver fibrosis/disease surrogates APRI, FIB4, LSM and the ©LiverRisk score (Figs. 4, S12-S14).

**Fig. 3.**
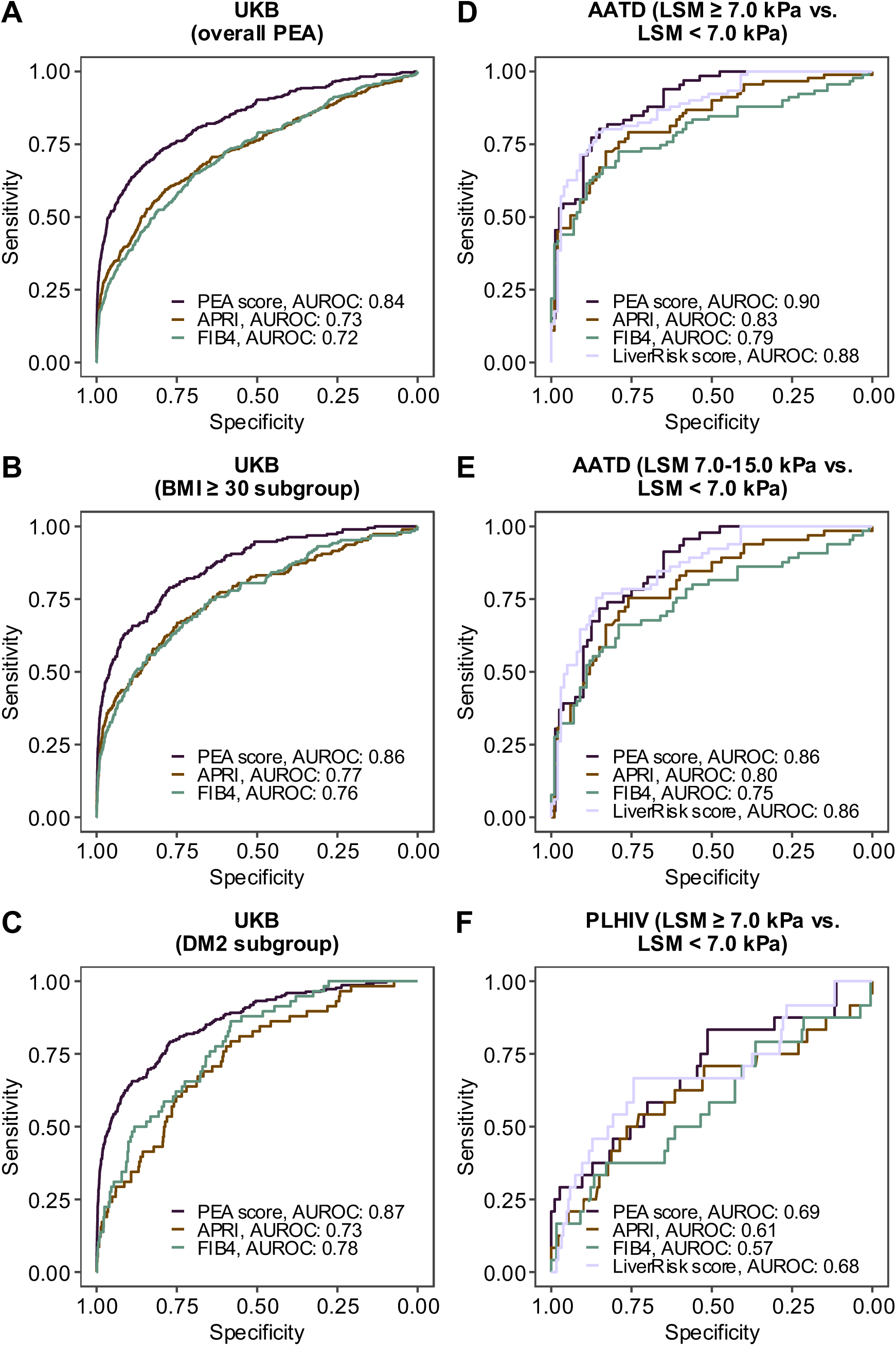
Comparison of a proximity extension assay (PEA) score with established liver fibrosis indices across patient cohorts. Receiver-operating curves (ROCs) displaying the performance of a PEA score in comparison to AST-to-platelet-ratio index (APRI), Fibrosis-4 index (FIB4) and the ©LiverRisk score. **A-C:** Prediction of major adverse liver outcomes (MALOs) in the overall UK Biobank (UKB) population with available PEA-data [A], and subgroups of UKB patients with obesity (BMI ≥30 kg/m²) [B] or type-2 diabetes (DM2) [C]. **D-E:** Differentiation between patients with alpha1-antitrypsin deficiency (AATD) based on liver stiffness measurements (LSM) via FibroScan®. Patients with significant liver fibrosis (D: LSM ≥7.0 kPa, E: LSM between 7.0-15.0 kPa, *i.e.,* excluding patients with possible clinically significant portal hypertension) were compared against those without (LSM <7.0 kPa). **F:** LSM-based differentiation between people living with HIV (PLHIV) with (LSM ≥7.0 kPa) and without significant liver fibrosis (LSM <7.0 kPa). The areas under receiver-operating curve (AUROC) are shown.

**Fig. 4.**
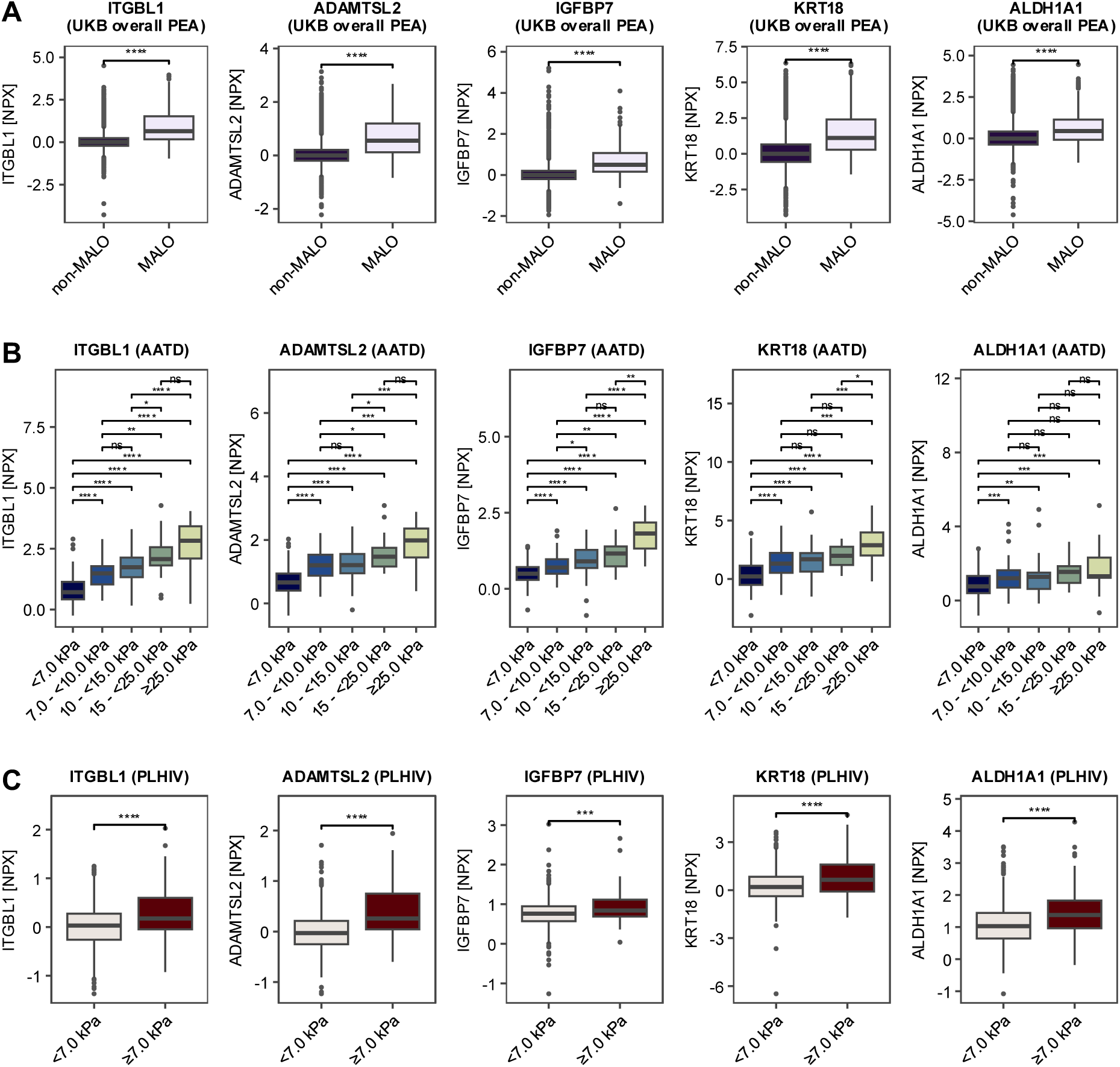
Analysis of the identified proteomic markers associated with significant liver disease across different patient populations. **A**: Plasma levels (normalised protein expression [NPX] values) of five protein markers (ITGBL1, ADAMTSL2, IGFBP7, KRT18, and ALDH1A1) in the overall UK Biobank (UKB) population with available proximity extension assay (PEA)-based proteomic data. Baseline measurements from patients with (MALO) and without major adverse liver outcomes during follow-up (non-MALO) are compared. **B**: Plasma levels of the above-described biomarkers in patients with alpha1-antitrypsin deficiency (AATD) with different liver fibrosis stages as indicated by liver stiffness measurement (LSM) (<7.0 kPa, 7.0-10.0 kPa, 10.0-15.0 kPa, 15.0-25.0 kPa, and ≥25.0 kPa). **C**: Plasma levels of the above-described biomarkers in people living with HIV (PLHIV) with vs. without LSM-based significant fibrosis (*i.e.,* LSM ≥7.0 vs. <7.0 kPa). Box plots indicate the median (central line), 25^th^-75^th^ percentiles (box), and smallest/largest non-outlier values (whiskers); outliers are depicted as black dots. Significance levels are indicated as follows: \**p* <0.05, \*\**p* <0.01, \*\*\**p* <0.001, \*\*\*\**p* <0.0001 (Wilcoxon rank-sum test). *ADAMTSL2: ADAMTS-like protein 2; ALDH1A1: aldehyde dehydrogenase 1A1; IGFBP7: insulin-like growth factor-binding protein 7; ITGBL1: integrin beta-like protein 1; KRT18: keratin-18*.

**Table 4:**
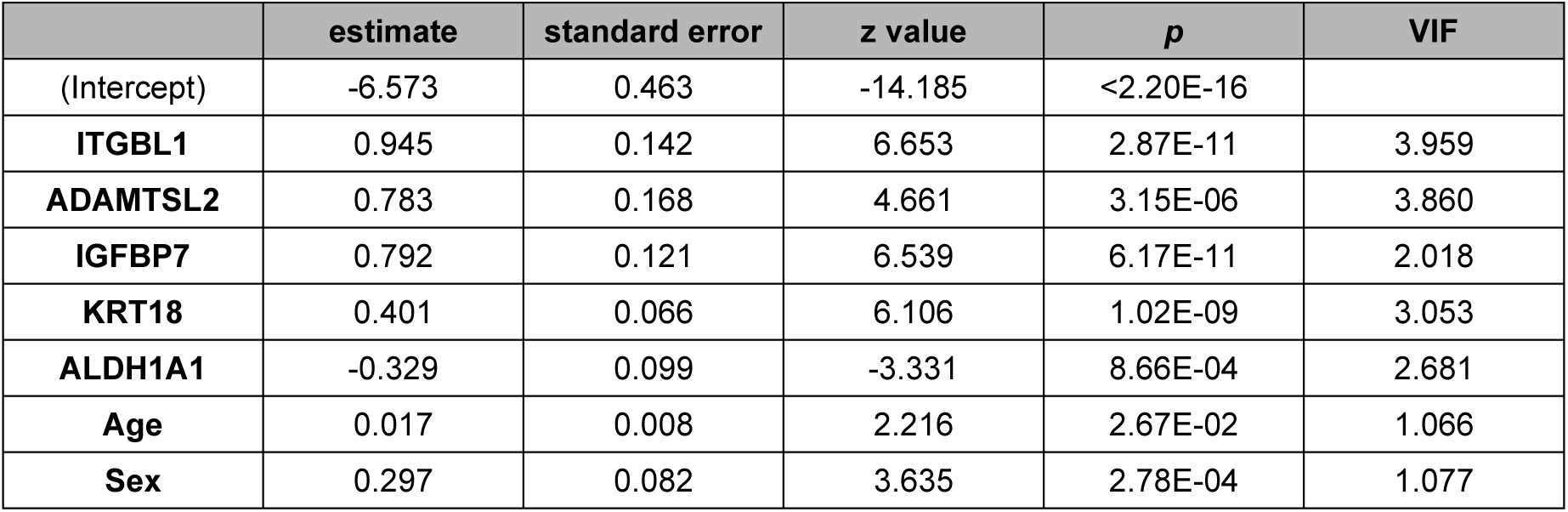
Composition of the proximity extension assay (PEA) score. ADAMTSL2: ADAMTS-like protein 2; ALDH1A1: aldehyde dehydrogenase 1A1; IGFBP7: insulin-like growth factor-binding protein 7; ITGBL1: integrin beta-like protein 1; KRT18: keratin-18; VIF: variance inflation factor.

|  | estimate | standard error | z value | p | VIF |
| --- | --- | --- | --- | --- | --- |
| (Intercept) | -6.573 | 0.463 | -14.185 | <2.20E-16 |  |
| **ITGBL1** | 0.945 | 0.142 | 6.653 | 2.87E-11 | 3.959 |
| **ADAMTSL2** | 0.783 | 0.168 | 4.661 | 3.15E-06 | 3.860 |
| **IGFBP7** | 0.792 | 0.121 | 6.539 | 6.17E-11 | 2.018 |
| **KRT18** | 0.401 | 0.066 | 6.106 | 1.02E-09 | 3.053 |
| **ALDH1A1** | -0.329 | 0.099 | -3.331 | 8.66E-04 | 2.681 |
| **Age** | 0.017 | 0.008 | 2.216 | 2.67E-02 | 1.066 |
| **Sex** | 0.297 | 0.082 | 3.635 | 2.78E-04 | 1.077 |

Collectively, our data indicate that PEA reliably quantifies several liver-related parameters and that PEA-based assessment can be used to develop prognostically useful liver scores.

## Discussion

The study objective was to evaluate the usefulness of the emerging PEA technique as a source of liver-related biomarkers. First, we demonstrated that analyses of three different cohorts yielded similar top biomarkers even though they have been performed in different PEA facilities, which confirms the robustness and standardisation of the assay. In line with that, the levels of several parameters measured by PEA strongly correlated with values obtained by alternative techniques. However, this may not be the case for all biomarkers[25], and because of that, the markers of interest need to be individually validated. Second, we identified 20 biomarkers that were consistently altered in all assessed cohorts and used a bioinformatic analysis to obtain a novel five-feature score. This comprised three HSC-related and two epithelial proteins. Notably, the HSC-based markers were previously reported as attractive liver fibrosis surrogates[6,10,26–30]. In addition, KRT18 is frequently assessed by the well-established M30/M65-ELISAs and is considered a marker of histological disease severity[26–28]. ALDH1A1, as the second epithelial protein is strongly expressed in pericentral hepatocytes and might therefore be particularly useful to detect an injury in this compartment[6,29–31]. The fact that these five biomarkers emerged from an unbiased analysis of ∼3,000 proteins strongly supports the importance of HSC and epithelial surrogates as prognosticators of liver disease severity. This is in line with data from a different affinity-based proteomic technique as well as mass spectrometry-based analysis, which identified similar proteomic signatures[6,10,32,33]. Notably, the 20 consistently altered proteins also contained several biomarkers linked to inflammation and endothelial function. The presence of an endothelial-related signature is not surprising, as endothelial dysfunction is important for liver disease progression and the development of portal hypertension, but might be less relevant in earlier disease stages[34,35]. Similarly, the identification of the Kupffer cell-related markers is well in line with their importance in both liver injury and homeostasis[36]. Further studies are needed to test whether the addition of these representatives provides further prognostic benefits.

Our analysis has several strengths. First, all assessed cohorts included a systematic evaluation of key liver-related parameters (*i.e.,* liver fibrosis or MALO) that are strongly related to liver-related death[23]. Second, while metabolic liver disease is the prevailing liver disease aetiology in the UKB[37], the results were validated in a cohort with a genetically determined, proteotoxic liver injury[15] as well as an HIV-related cohort[16]. To the best of our knowledge, our study is the first one using a large-scale serum proteomics to analyse biomarkers of AATD-associated liver disease. This is of obvious importance given that AATD-associated liver disease constitutes an area of active drug development[38] and our findings may support these efforts. The strong overlap between biomarkers seen in UKB and the AATD cohort suggests that they are aetiology-independent, which meshes well with their ability to reflect essential fibrosis-related processes such as HSC activation. Another strength of the study is the fact that we were able to prognosticate the development of MALOs, which is difficult to study due to its uncommonness, as well as the slow rate of liver fibrosis progression[39,40]. At the same time, we were able to evaluate the usefulness of the detected biomarkers/the designed score in the PLHIV cohort, consisting primarily of subjects with no/minimal fibrosis, as well as three high-risk cohorts (*i.e.,* diabetes, obesity and AATD). Finally, the described score outperformed the routinely used APRI/FIB4 scores and was similar or even slightly better than the recently described ©LiverRisk score. The somewhat lower performance of the score in the PLHIV cohort might be because this cohort assessed primarily subjects with minimal-to-moderate fibrosis, and the LSM that was used as a comparator has a suboptimal discriminative ability in this range[41]. Compared to other proteomic approaches, our PEA score achieved performance comparable or slightly higher than scores described for the SomaScan technique, despite using fewer biomarkers[42]. However, head-to-head studies are needed to compare both techniques.

Despite the above-described strengths, our study also has limitations. Most participants in both cohorts are of European descent[20,43], and future studies should therefore validate our findings in other ethnicities. An analysis of additional longitudinal cohorts with enough MALOs would also be desirable. Finally, our analyses focused primarily on hepatocellular liver disorders, and the applicability of our score to cholestatic diseases remains to be tested.

In conclusion, our study demonstrates the usefulness of a new PEA proteomic technique to determine the liver fibrosis stage and to prognosticate the development of MALOs. Given that this non-invasive approach requires only a small sample volume, is standardised across facilities, and quantifies proteins across a large dynamic range of protein concentrations [7], it may become a useful tool for both patient stratification and drug development.

## Data Availability

Data is available upon reasonable request to the corresponding author.

## Abbreviations

AATD: alpha1-antitrypsin deficiency
ADAMTSL2: ADAMTS-like protein 2
ALDH1A1: aldehyde dehydrogenase 1A1
ALT: alanine aminotransferase
APRI: AST-to-platelet-ratio index
AST: aspartate aminotransferase
AUROC: area under receiver-operating curve
DM2: type-2 diabetes
FDR: false discovery rate
FIB4: Fibrosis-4 index
GGT: gamma-glutamyltransferase
HSC: hepatic stellate cell
ICD-10: International Classification of Diseases version 10
IGFBP7: insulin-like growth factor-binding protein 7
ITGBL1: integrin beta-like protein 1
KRT18: keratin-18
LSM: liver stiffness measurement
MALO: major adverse liver outcomes
MENT, C1orf56: chromosome 1 open reading frame 56
NPX: normalised protein expression
OPCS-4: operations/procedure codes version 4
PLHIV: people living with HIV
PEA: proximity extension assay
Pi*ZZ: homozygous Pi*Z mutation in alpha1-antitrypsin gene
TE: transient elastography
THBS2: thrombospondin-2
UKB: the UK Biobank cohort.

## Authorship contribution statement

Study concept and design: K.R., P.S.

Acquisition of data: S.B., K.R., AS.K, C.S., M.F., L.E.v.E., L.A.B.J.

Analysis and interpretation of data: S.B., AS.K., K.R., P.T.

Drafting of the manuscript: S.B., K.R., P.S.

Critical revision of the manuscript for important intellectual content: K.R., P.S.

Figures and tables: S.B., J.A.B.

Statistical analysis: S.B., A.S.K., K.R., P.T.

Obtained funding: P.S., C.K., B.Z., M.L., P.K.

Administrative, technical or material support: C.S., M.F., C.K., B.Z., M.L., P.K., L.E.v.E., L.A.B.J.

Study supervision: K.R., P.S.

All authors had full access to all the data and approved the final version of this manuscript. All authors take responsibility for the integrity of the data and the accuracy of the data analysis.

