## Supplement for "Affinity proteomics-based non-invasive detection of clinically significant liver disease"

### Supplementary Methods

#### Statistical analyses

All analyses were performed using the R environment[1](R Foundation, Vienna, Austria, version 4.4.1) and R Studio[2](version 2024.04.2+764) along with following R packages: dplyr[3], ggplot2[4], cowplot[5], limma[6], SummarizedExperiment[7] and pROC[8].

For proteomic analyses, differentially abundant proteins were identified using linear models with empirical Bayes moderation, with age and sex included as covariates. P-values were adjusted for multiple testing using the false discovery rate (FDR) method. Proteins were considered significantly differentially abundant if their FDR was <0.05 across a given condition.

Proteins of interest were mapped to both publicly available tissue expression datasets (accessible at https://www.proteinatlas.org/humanproteome/tissue) and liver single-cell RNA sequencing data ([9], accessed via www.livercellatlas.org).

For the comparison of protein levels between two groups, Wilcoxon rank sum tests were used. Box plots were used to visualise protein levels across different groups. Correlations between selected variables were assessed in a pairwise manner using Spearman's rank correlation test.

Demographic and clinical parameters were compared using linear regression models. When appropriate, models were adjusted for age, sex and BMI.

AST-to-platelet-ratio index (APRI) and Fibrosis-4 index (FIB4) were calculated using the following formulas: APRI = ((AST[IU/L]/34.8)/platelet count[x$10^9$/L]); FIB4 = (age x AST[IU/L]/platelet count[x$10^9$/L] x √ALT[IU/L]); and used with previously established cut-offs for significant/advanced liver fibrosis in figures[10–12]. The ©LiverRisk score was calculated as discussed previously and used with established cut-offs[13].

Associations between proteomic biomarkers and mortality outcomes were studied via logistic regression. For each protein of interest, a separate model was constructed to evaluate the relationship with both overall and liver-related mortality (defined by International Classification of Diseases version 10 (ICD-10) codes K70-K77 as the primary cause of death). All models were adjusted for age, sex, and BMI as covariates. Results are presented as odds ratios (ORs) with 95% confidence intervals per standard deviation increase in plasma levels. To ensure comparability across biomarkers with different expression ranges, protein levels were standardised prior to the analysis.

**Development of prognostic score**

To develop a prognostic score, we first assessed the ability of parameters to discriminate between patients developing a major adverse liver outcome (MALO) and those without via logistic regression. All models included age and sex as covariates. For the modelling process, missing values were imputed using a modified k-nearest neighbour approach accounting for truncation at a minimum threshold (i.e., limit of detection)[14]. The subset of 20 proteomic variables, representing commonly altered proteins between the MALO-associated signature in the UK Biobank (UKB) and fibrosis-associated signatures in the AATD and PLHIV cohorts, were evaluated. Any feature reaching an area under receiver-operating curve (AUROC) >0.55 and p-value <0.05 in a univariable analysis was used. All models were analysed for collinearity, and variables with correlations greater than 0.7 or less than -0.7 were not included in the same model to mitigate collinearity and increase the stability and accuracy of estimated coefficients. Models were considered if all coefficients presented a significant association ($p$ <0.05) with MALO, and their performance was assessed with AUROCs. For quality assessment, variable inflation factors were assessed for the selected model. Goodness of fit was assessed using the Hosmer-Lemeshow test. The linearity between assessed variables and the logit of a future MALO was assessed both graphically and using the Box-Tidwell test.

### Supplementary Figures

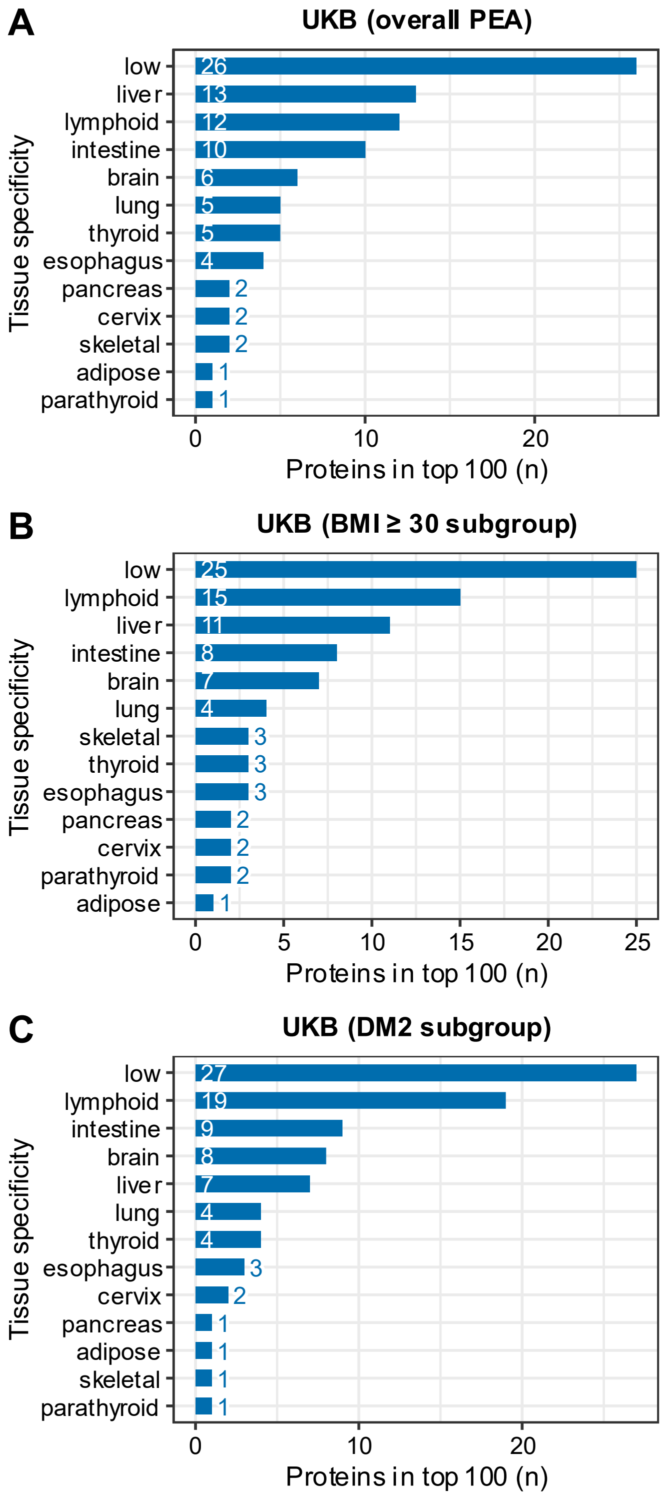

**Fig. S1. Mapping of the top 100 differentially abundant proteins in the United Kingdom Biobank (UKB) cohort to their tissue of origin.** Proteins that discriminate between subjects with/without future major adverse liver outcomes (MALOs) were assessed. The plots show the absolute number of proteins mapped to a specific tissue in (A) the entire proteomic cohort, (B) a subgroup of obese (BMI ≥30) patients and (C) a subgroup with type-2 diabetes (DM2). A single protein may be specific to more than one tissue. 20 (in A/C) and 23 (in B) proteins could not be assigned and are not shown in the plots above.

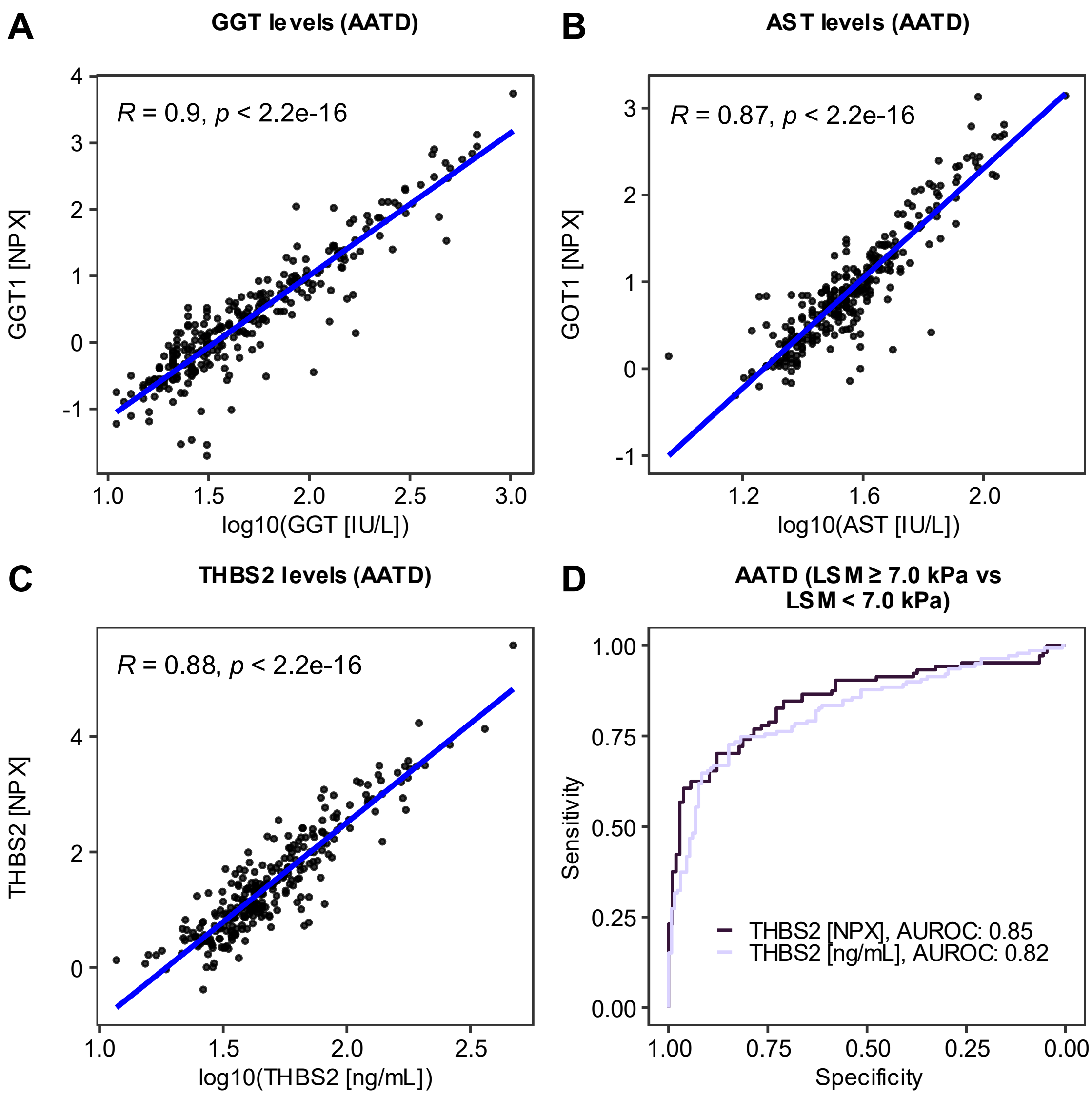

**Fig. S2: Comparing serum and proteomic measurements of biomarkers in the cohort of severe alpha1-antitrypsin deficiency (AATD) subjects.** A/B: Scatter plot depicts correlation of routine log10-transformed (in IU/L) and corresponding proximity extension assay (PEA)-based measurements of gamma-glutamyltransferase (GGT/GGT1) and aspartate aminotransferase (AST/GOT1) serum levels. C: Depicts the correlation between log10-transformed serum concentrations of thrombospondin-2 (THBS2, in ng/mL) determined via immunoassay (x-axis) and corresponding PEA measurements (Olink® platform, y-axis, in normalised protein expression [NPX] values). Blue lines represent the linear regression fit. Each dot represents an individual sample. Spearman rank correlation factors and p-values are depicted. D: Receiver operating curves (ROCs) display the ability of a PEA- and immunoassay-based THBS2 levels (in ng/mL) to distinguish AATD subjects with vs. without significant liver fibrosis assessed through liver stiffness measurements (LSM) via FibroScan®. AATD subjects with LSM ≥7.0 kPa and LSM <7.0 kPa were compared. The area under the receiver operating curve (AUROC) is shown.

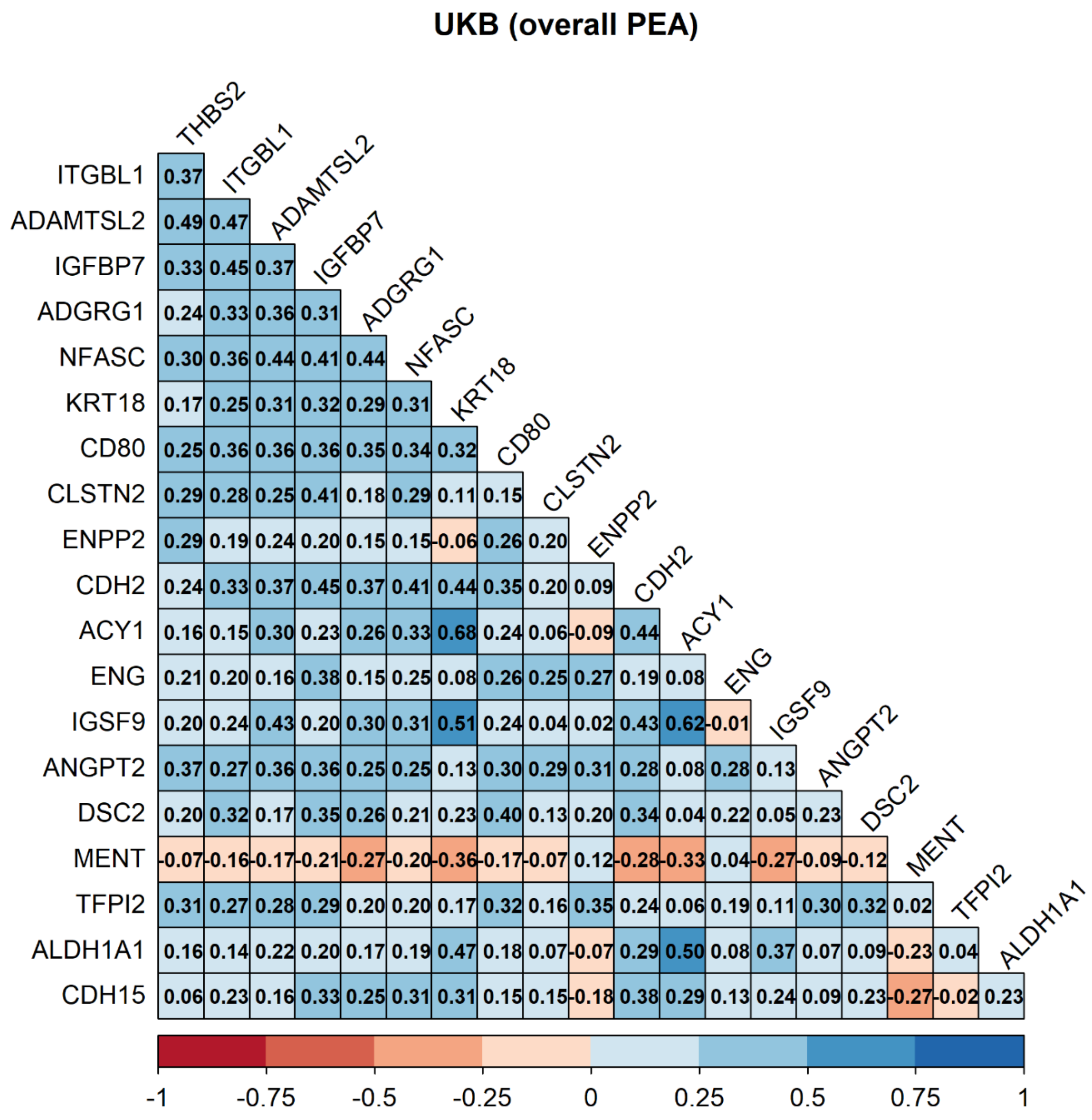

**Fig. S3: Correlation of 20 biomarkers in the UK Biobank (UKB) cohort with available proximity extension assay (PEA) proteomic data.** Displayed are the Spearman rank correlation coefficients. *ACY1: aminoacylase 1; ADAMTSL2: ADAMTS-like protein 2; ADGRG1: adhesion G protein-coupled receptor G1; ALDH1A1: aldehyde dehydrogenase 1A1; ANGPT2: angiopoietin 2; CD80: CD80 molecule; CDH2: cadherin 2; CDH15: cadherin 15; CLSTN2: calsyntenin 2; DSC2: desmocollin 2; ENG: endoglin; ENPP2: ectonucleotide pyrophosphatase/phosphodiesterase 2; IGFBP7: insulin-like growth factor-binding protein 7; IGSF9: Immunoglobulin superfamily member 9; ITGBL1: integrin beta-like protein 1; KRT18: keratin-18; MENT: C1orf56 (chromosome 1 open reading frame 56); NFASC: neurofascin; TFPI2: tissue factor pathway inhibitor 2; THBS2: thrombospondin-2.*

### UKB (BMI ≥ 30)

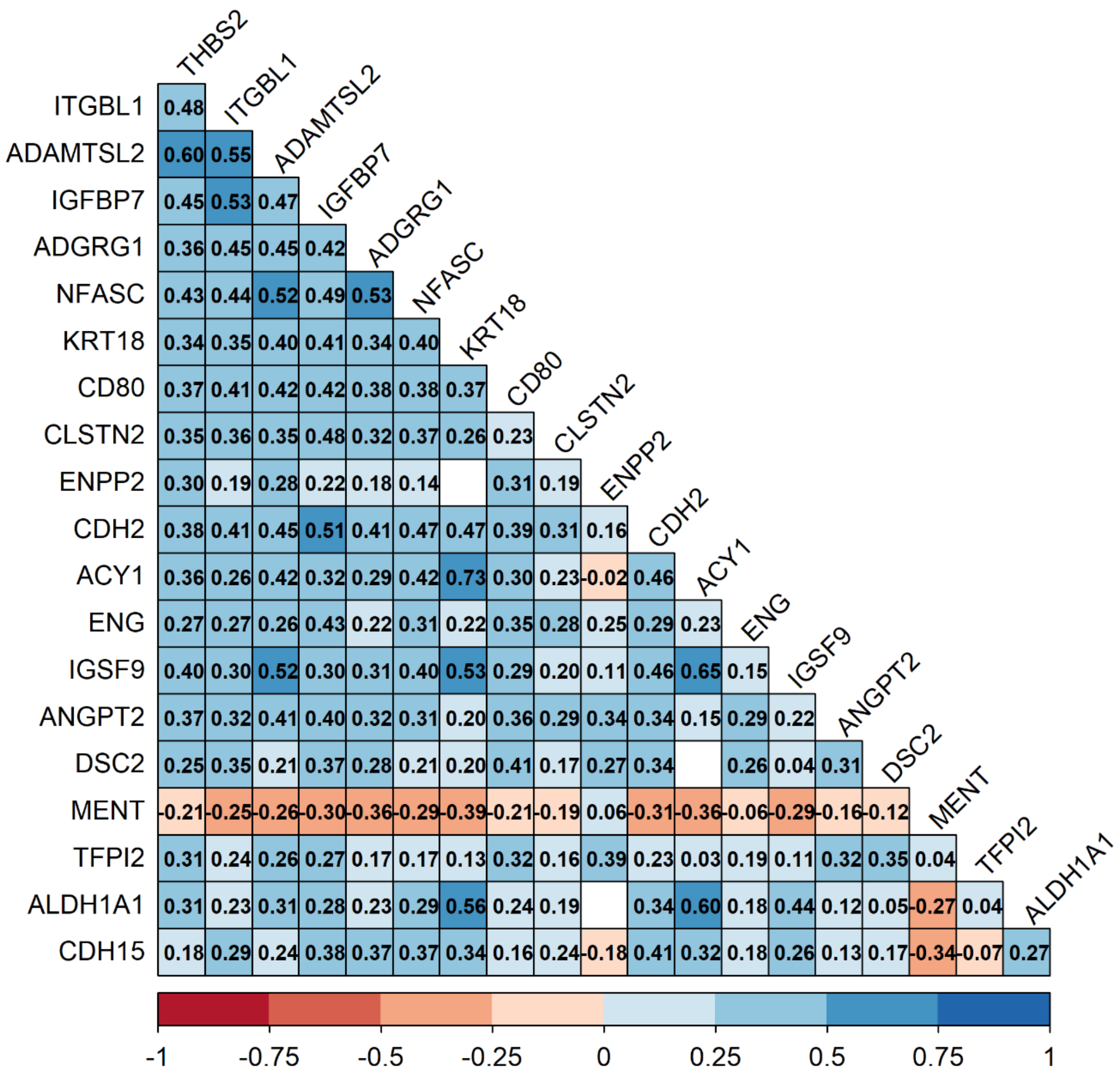

**Fig. S4: Correlation of 20 biomarkers in a subgroup of obese (BMI ≥30) UK Biobank (UKB) participants.** Displayed are the Spearman rank correlation coefficients. *ACY1: aminoacylase 1; ADAMTSL2: ADAMTS-like protein 2; ADGRG1: adhesion G protein-coupled receptor G1; ALDH1A1: aldehyde dehydrogenase 1A1; ANGPT2: angiopoietin 2; CD80: CD80 molecule; CDH2: cadherin 2; CDH15: cadherin 15; CLSTN2: calsyntenin 2; DSC2: desmocollin 2; ENG: endoglin; ENPP2: ectonucleotide pyrophosphatase/phosphodiesterase 2; IGFBP7: insulin-like growth factor-binding protein 7; IGSF9: Immunoglobulin superfamily member 9; ITGBL1: integrin beta-like protein 1; KRT18: keratin-18; NFASC: neurofascin; MENT: C1orf56 (chromosome 1 open reading frame 56); TFPI2: tissue factor pathway inhibitor 2; THBS2: thrombospondin-2.*

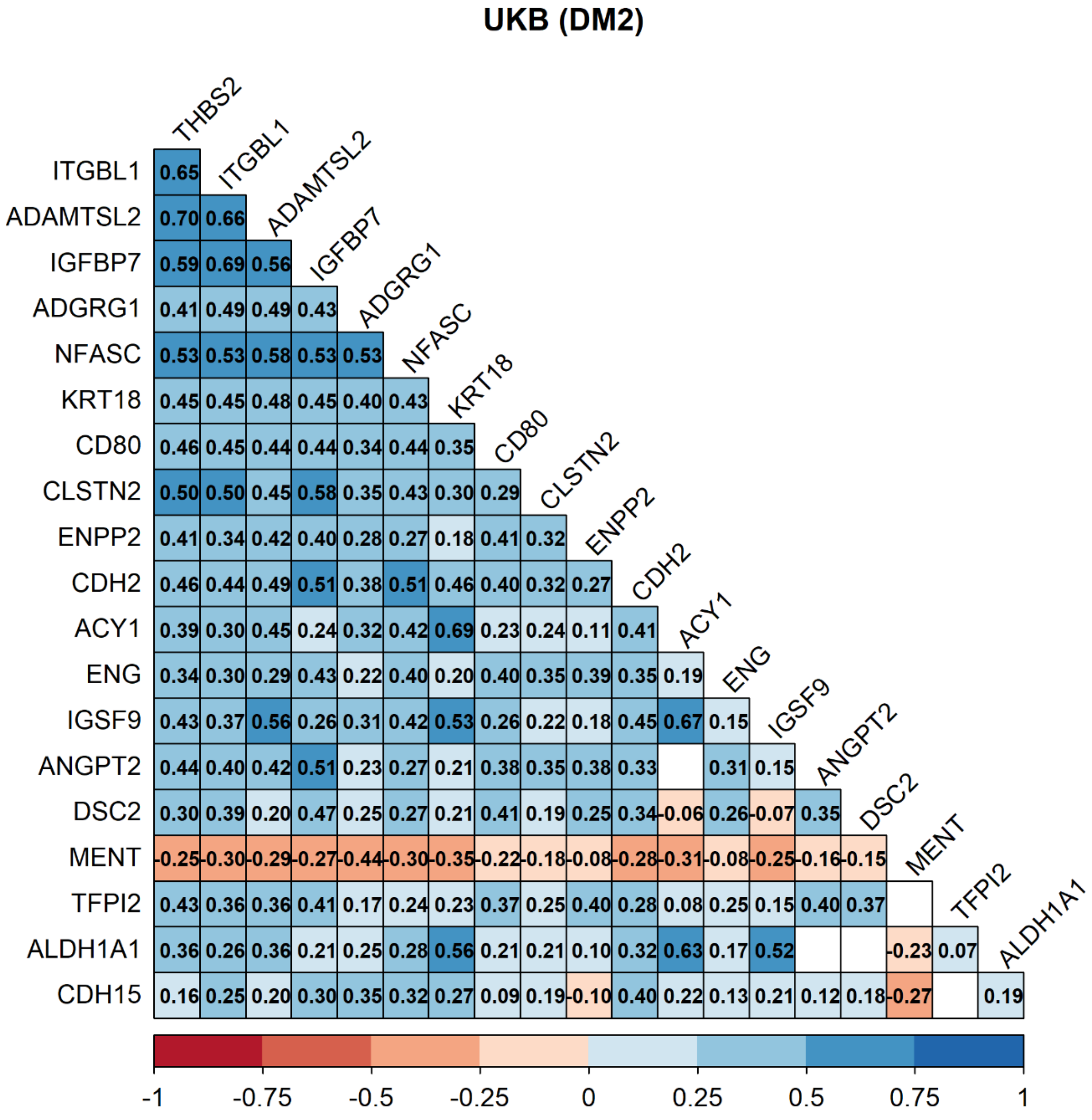

**Fig. S5: Correlation of 20 biomarkers in a subgroup of type-2 diabetic (DM2) UK Biobank (UKB) participants.** Displayed are the Spearman rank correlation coefficients. *ACY1: aminoacylase 1; ADAMTSL2: ADAMTS-like protein 2; ADGRG1: adhesion G protein-coupled receptor G1; ALDH1A1: aldehyde dehydrogenase 1A1; ANGPT2: angiopoietin 2; CD80: CD80 molecule; CDH2: cadherin 2; CDH15: cadherin 15; CLSTN2: calsyntenin 2; DSC2: desmocollin 2; ENG: endoglin; ENPP2: ectonucleotide pyrophosphatase/phosphodiesterase 2; IGFBP7: insulin-like growth factor-binding protein 7; IGSF9: Immunoglobulin superfamily member 9; ITGBL1: integrin beta-like protein 1; KRT18: keratin-18; MENT: C1orf56 (chromosome 1 open reading frame 56); NFASC: neurofascin; TFPI2: tissue factor pathway inhibitor 2; THBS2: thrombospondin-2.*

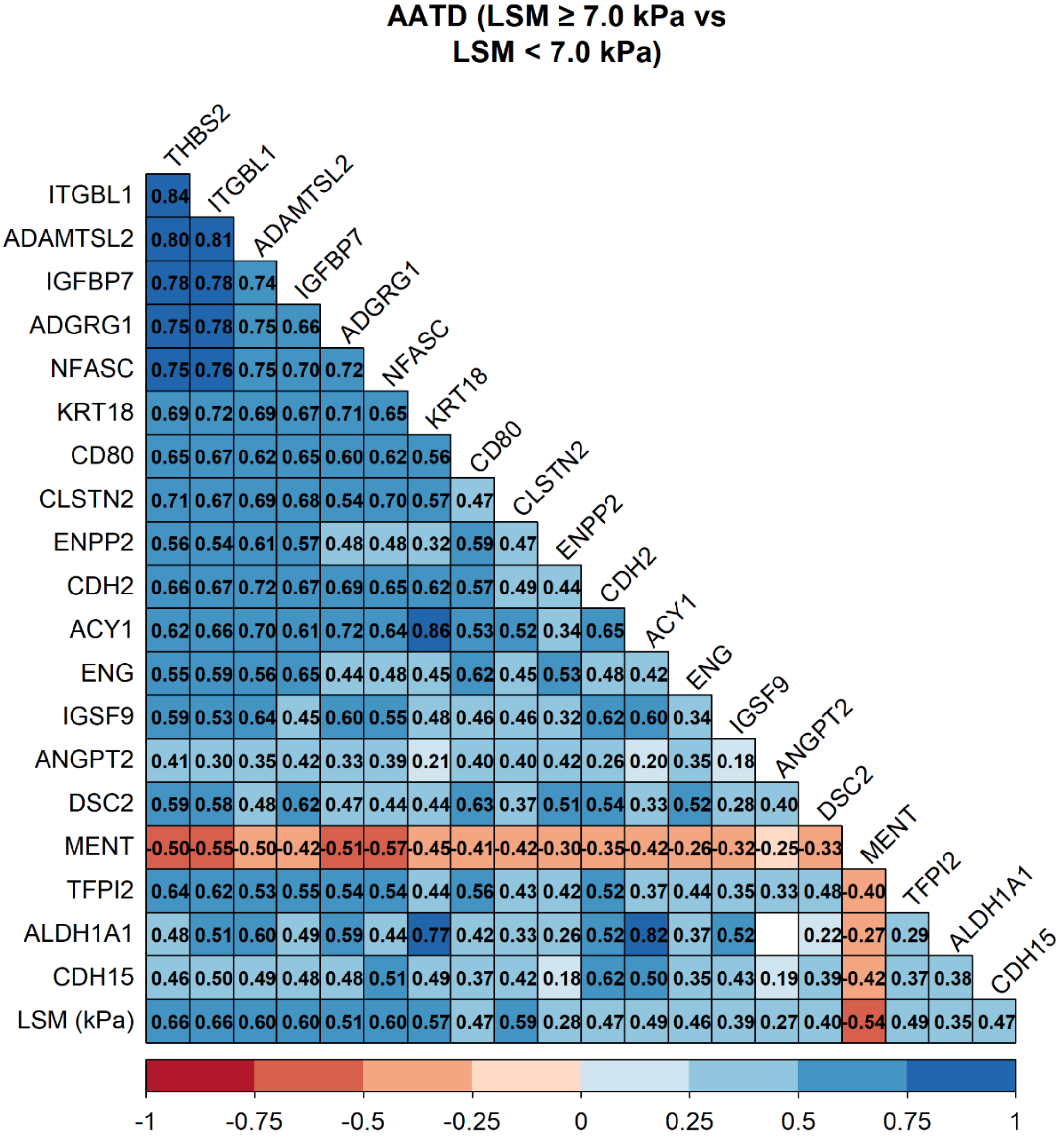

**Fig. S6: Correlation of 20 biomarkers in the cohort of alpha1-antitrypsin deficiency (AATD) subjects.** Displayed are the Spearman rank correlation coefficients. Liver fibrosis was assessed with non-invasive liver stiffness measurement (LSM, reported in kPa, via FibroScan®). *ACY1: aminoacylase 1; ADAMTSL2: ADAMTS-like protein 2; ADGRG1: adhesion G protein-coupled receptor G1; ALDH1A1: aldehyde dehydrogenase 1A1; ANGPT2: angiopoietin 2; CD80: CD80 molecule; CDH2: cadherin 2; CDH15: cadherin 15; CLSTN2: calsyntenin 2; DSC2: desmocollin 2; ENG: endoglin; ENPP2: ectonucleotide pyrophosphatase/phosphodiesterase 2; IGFBP7: insulin-like growth factor-binding protein 7; IGSF9: Immunoglobulin superfamily member 9; ITGBL1: integrin beta-like protein 1; KRT18: keratin-18; MENT: C1orf56 (chromosome 1 open reading frame 56); NFASC: neurofascin; TFPI2: tissue factor pathway inhibitor 2; THBS2: thrombospondin-2.*

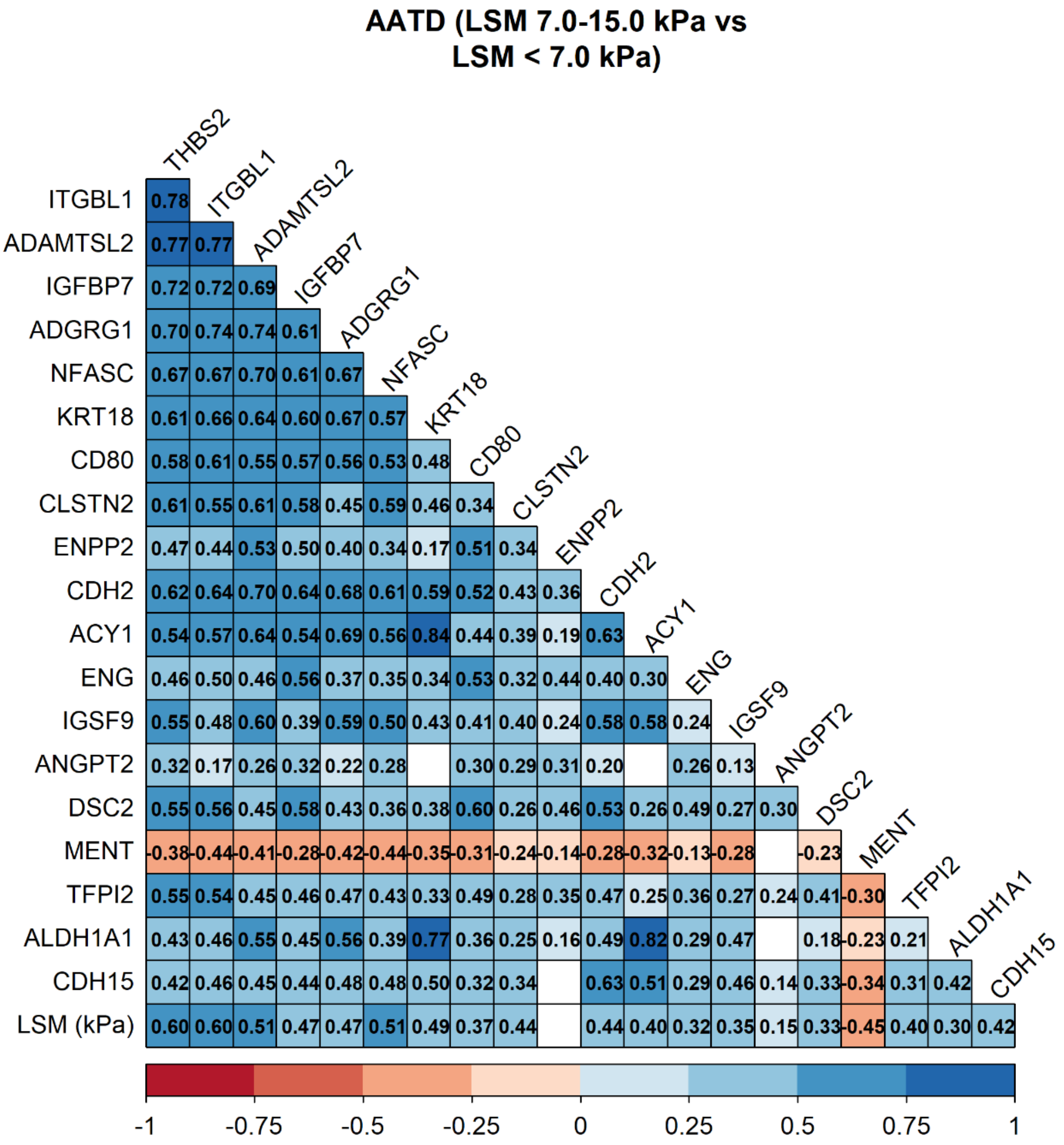

**Fig. S7: Correlation of 20 biomarkers in a subcohort of alpha1-antitrypsin deficiency (AATD) subjects without possible clinically significant portal hypertension.** Displayed are the Spearman rank correlation coefficients. Liver fibrosis was assessed with non-invasive liver stiffness measurement (LSM, reported in kPa, via FibroScan®) and subjects with LSM ≥15kPa were excluded. *ACY1: aminoacylase 1; ADAMTSL2: ADAMTS-like protein 2; ADGRG1: adhesion G protein-coupled receptor G1; ALDH1A1: aldehyde dehydrogenase 1A1; ANGPT2: angiopoietin 2; CD80: CD80 molecule; CDH2: cadherin 2; CDH15: cadherin 15; CLSTN2: calsyntenin 2; DSC2: desmocollin 2; ENG: endoglin; ENPP2: ectonucleotide pyrophosphatase/phosphodiesterase 2; IGFBP7: insulin-like growth factor-binding protein 7; IGSF9: Immunoglobulin superfamily member 9; ITGBL1: integrin beta-like protein 1; KRT18: keratin-18; MENT: C1orf56 (chromosome 1 open reading frame 56); NFASC: neurofascin; TFPI2: tissue factor pathway inhibitor 2; THBS2: thrombospondin-2.*

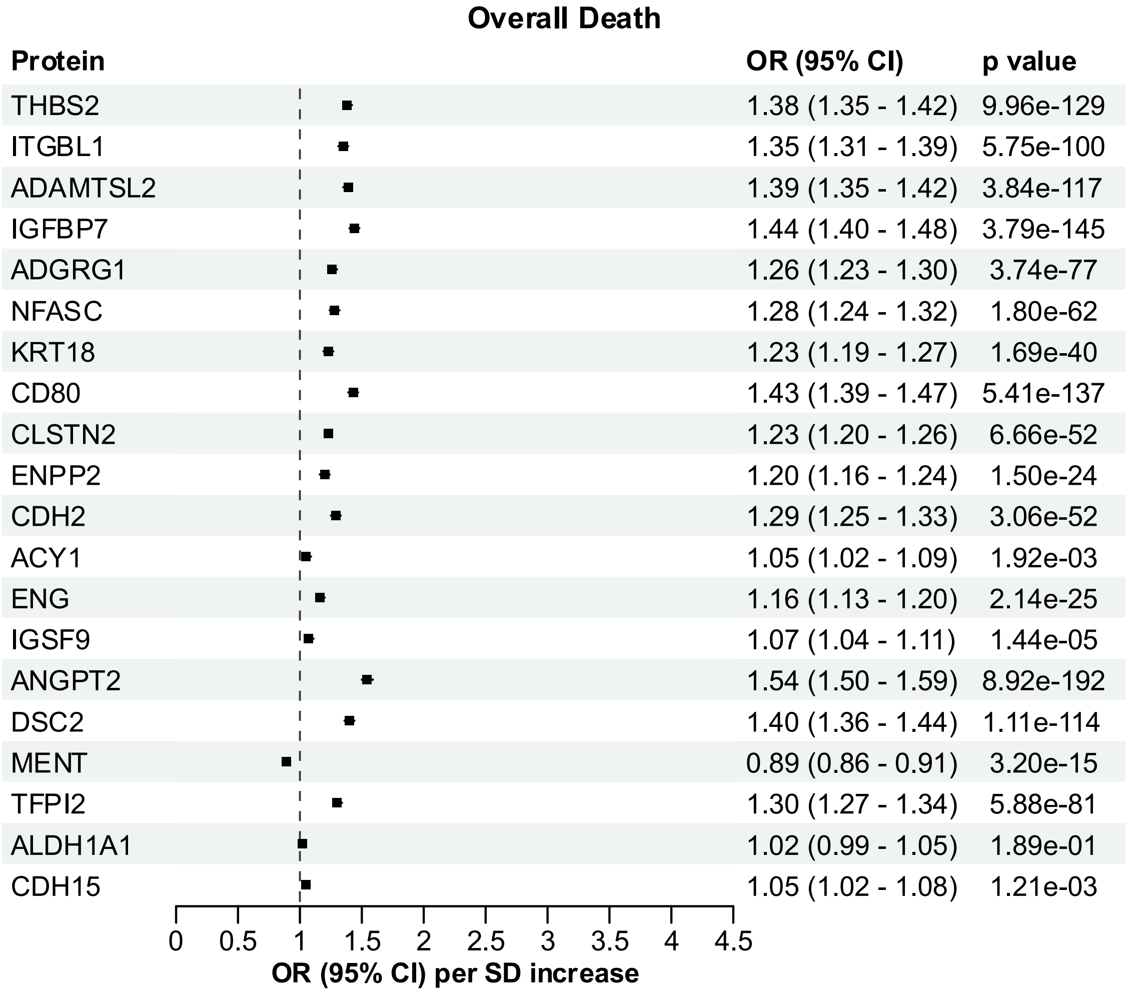

**Fig. S8. Association of overall mortality with plasma levels of selected proteomic biomarkers in the UK Biobank (UKB).** Odds ratios (ORs) and p-values were generated from logistic regression models (adjusted for age, sex, and BMI). ORs (95% CI) per standard deviation increase of plasma levels are shown. *ACY1: aminoacylase 1; ADAMTSL2: ADAMTS-like protein 2; ADGRG1: adhesion G protein-coupled receptor G1; ALDH1A1: aldehyde dehydrogenase 1A1; ANGPT2: angiopoietin 2; CD80: CD80 molecule; CDH2: cadherin 2; CDH15: cadherin 15; CLSTN2: calsyntenin 2; DSC2: desmocollin 2; ENG: endoglin; ENPP2: ectonucleotide pyrophosphatase/phosphodiesterase 2; IGFBP7: insulin-like growth factor-binding protein 7; IGSF9: Immunoglobulin superfamily member 9; ITGBL1: integrin beta-like protein 1; KRT18: keratin-18; MENT: C1orf56 (chromosome 1 open reading frame 56); NFASC: neurofascin; TFPI2: tissue factor pathway inhibitor 2; THBS2: thrombospondin-2.*

**Liver-related Death (K7)**

| Protein | OR (95% CI) | p value |
|---|---|---|
| THBS2 | 2.32 (2.11 - 2.56) | 1.66e-64 |
| ITGBL1 | 2.37 (2.14 - 2.62) | 3.06e-61 |
| ADAMTSL2 | 2.77 (2.47 - 3.12) | 4.64e-67 |
| IGFBP7 | 1.94 (1.77 - 2.13) | 9.58e-44 |
| ADGRG1 | 2.42 (2.10 - 2.78) | 3.84e-35 |
| NFASC | 3.40 (2.89 - 4.00) | 3.85e-49 |
| KRT18 | 2.65 (2.27 - 3.08) | 1.21e-36 |
| CD80 | 2.27 (2.02 - 2.56) | 3.52e-42 |
| CLSTN2 | 2.11 (1.88 - 2.35) | 1.62e-40 |
| ENPP2 | 2.68 (2.28 - 3.12) | 8.86e-35 |
| CDH2 | 3.26 (2.64 - 4.02) | 5.37e-28 |
| ACY1 | 3.44 (2.86 - 4.13) | 1.84e-39 |
| ENG | 2.48 (2.11 - 2.92) | 1.10e-27 |
| IGSF9 | 3.16 (2.62 - 3.79) | 1.03e-34 |
| ANGPT2 | 2.05 (1.73 - 2.41) | 7.82e-18 |
| DSC2 | 1.66 (1.42 - 1.93) | 4.96e-11 |
| MENT | 0.41 (0.35 - 0.48) | 4.99e-28 |
| TFPI2 | 1.37 (1.25 - 1.48) | 4.46e-13 |
| ALDH1A1 | 2.15 (1.83 - 2.50) | 5.72e-22 |
| CDH15 | 1.48 (1.23 - 1.76) | 1.71e-05 |

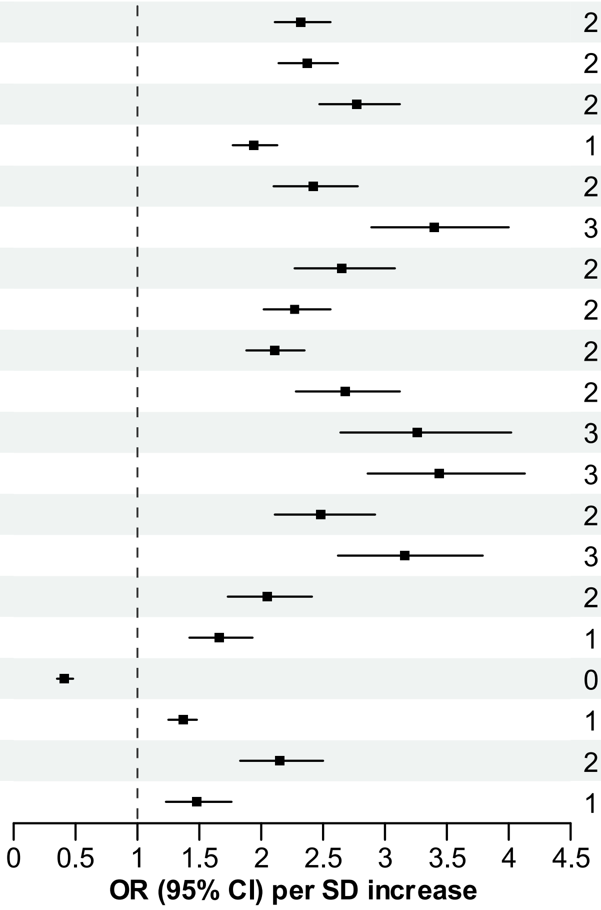

**Fig. S9. Association of liver-related mortality with plasma levels of selected proteomic biomarkers in the UK Biobank (UKB).** Odds ratios (ORs) and p-values were generated from logistic regression models (adjusted for age, sex, and BMI). ORs (95% CI) per standard deviation increase of plasma levels are shown. Liver-related mortality was defined via primary causes of death related to ICD-10 diagnosis codes K70 to K77. *ACY1: aminoacylase 1; ADAMTSL2: ADAMTS-like protein 2; ADGRG1: adhesion G protein-coupled receptor G1; ALDH1A1: aldehyde dehydrogenase 1A1; ANGPT2: angiopoietin 2; CD80: CD80 molecule; CDH2: cadherin 2; CDH15: cadherin 15; CLSTN2: calsyntenin 2; DSC2: desmocollin 2; ENG: endoglin; ENPP2: ectonucleotide pyrophosphatase/phosphodiesterase 2; ICD-10: International Classification of Diseases version 10; IGFBP7: insulin-like growth factor-binding protein 7; IGSF9: Immunoglobulin superfamily member 9; ITGBL1: integrin beta-like protein 1; KRT18: keratin-18; MENT: C1orf56 (chromosome 1 open reading frame 56); NFASC: neurofascin; TFPI2: tissue factor pathway inhibitor 2; THBS2: thrombospondin-2.*

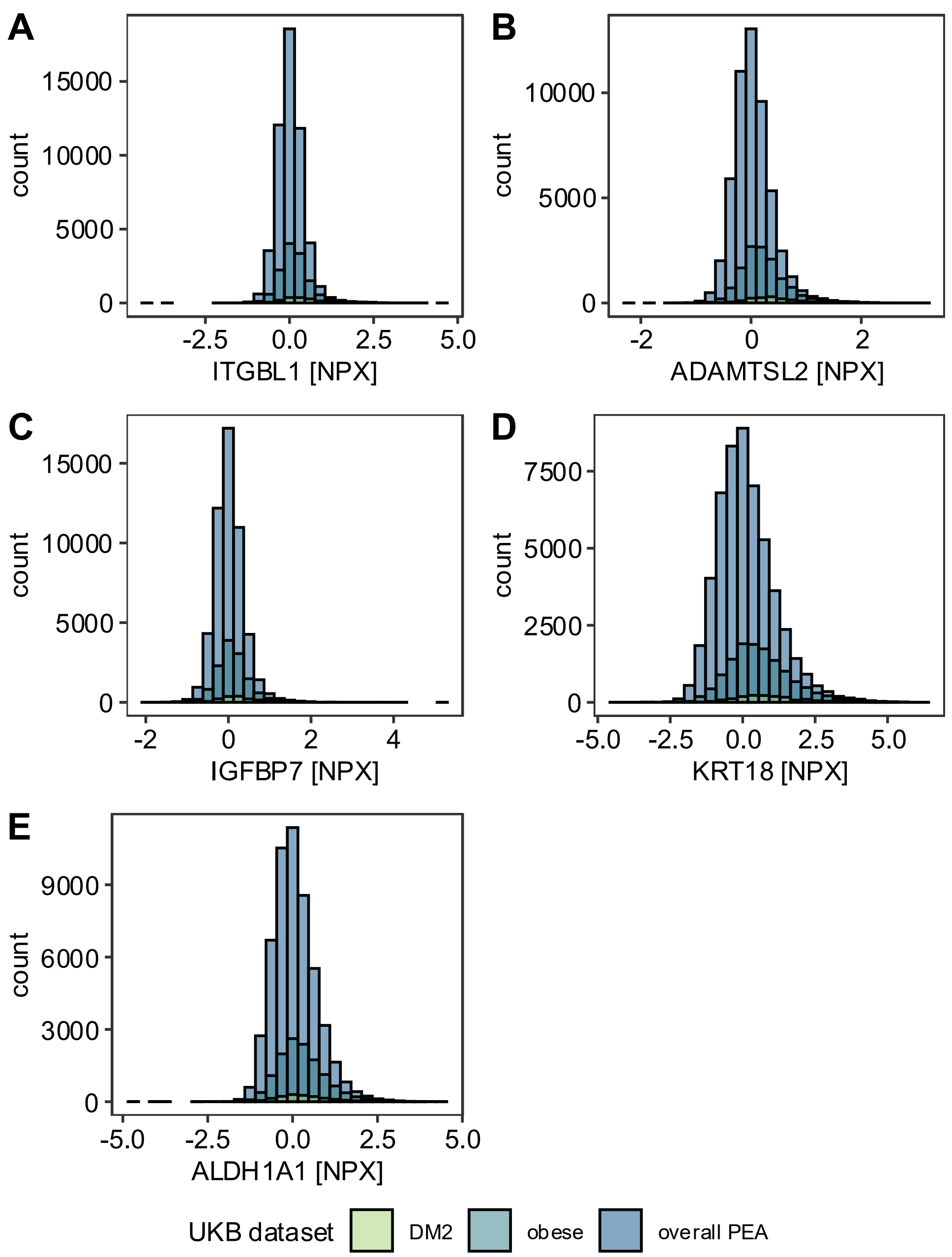

**Fig. S10: Distribution of continuous parameters assessed in multivariable logistic regression during development of PEA score.** Histograms display the distribution of the levels of the depicted five parameters in the UK Biobank (UKB) cohort (x-axis, normalised expression values [NPX]). A: integrin beta-like protein 1 [ITGBL1]; B: ADAMTS-like protein 2 [ADAMTSL2]; C: insulin-like growth factor-binding protein 7 [IGFBP7]; D: keratin-18 [KRT18]; E: aldehyde dehydrogenase 1A1 [ALDH1A1]. *DM2: type-2 diabetes; PEA: proximity extension assay.*

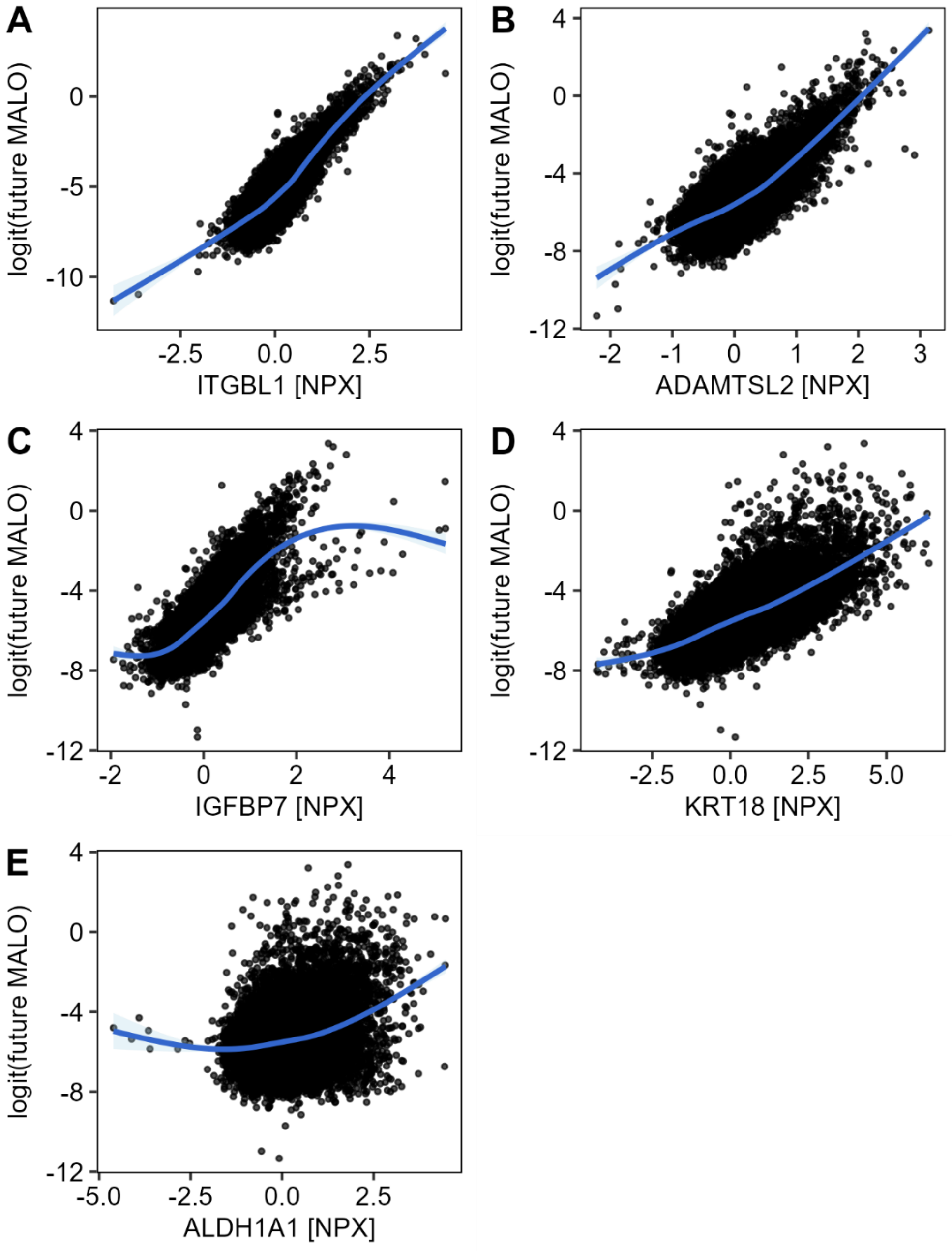

**Fig. S11: The relationship between parameters in the PEA score and the logit of the development of a future major adverse liver outcome (MALO).** Scatter plots visualise the relationship between the parameters chosen for the PEA score in the UK Biobank cohort (x-axis, normalised expression values [NPX]) and the logit of a future MALO (y-axis). A: integrin beta-like protein 1 [ITGBL1]; B: ADAMTS-like protein 2 [ADAMTSL2]; C: insulin-like growth factor-binding protein 7 [IGFBP7]; D: keratin-18 [KRT18]; E: aldehyde dehydrogenase 1A1 [ALDH1A1]. *PEA: proximity extension assay.*

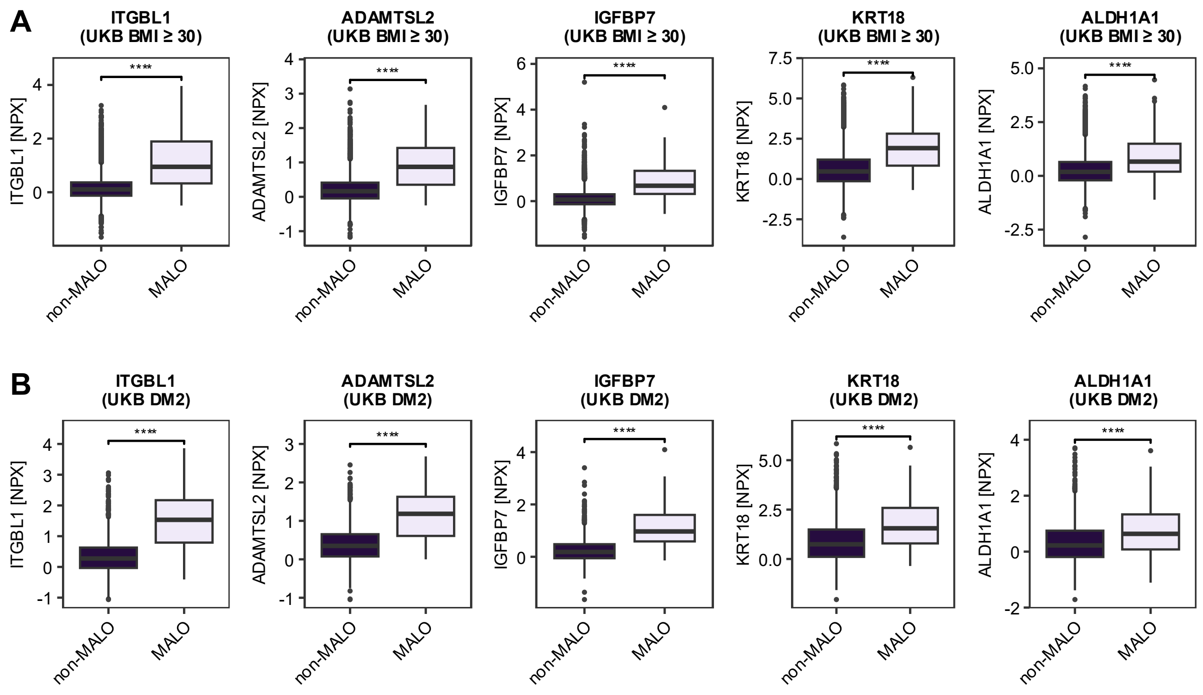

**Fig. S12: Levels of selected proteomic markers in obese and diabetic subjects from the UK Biobank (UKB) subdivided based on the development of major adverse liver outcomes (MALOs).** Box plots display relative expression levels (NPX values) of five protein markers (ITGBL1, ADAMTSL2, IGFBP7, KRT18, and ALDH1A1) in subgroups of UKB subjects with A: obesity (BMI ≥30) and B: type-2 diabetes (DM2), comparing subjects with versus those without MALOs. A Wilcoxon rank sum test was used to calculate p-values. Significance levels are indicated as follows: ns: not significant, *$p$ <0.05, **$p$ <0.01, ***$p$ <0.001, ****$p$ <0.0001. *ADAMTSL2: ADAMTS-like protein 2; ALDH1A1: aldehyde dehydrogenase 1A1; IGFBP7: insulin-like growth factor-binding protein 7; ITGBL1: integrin beta-like protein 1; KRT18: keratin-18.*

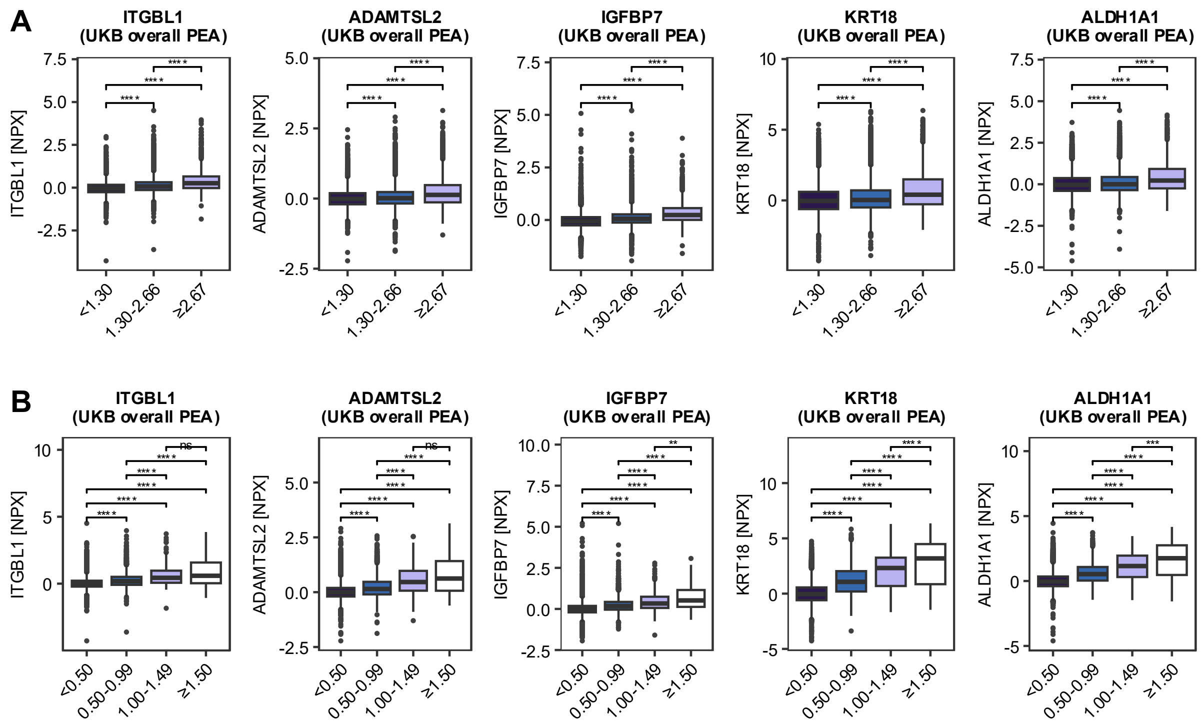

**Fig. S13: Ability of selected biomarkers to discriminate between liver fibrosis stages in the UK Biobank (UKB) cohort.** Box plots display relative expression levels (NPX values) of five protein markers (ITGBL1, ADAMTSL2, IGFBP7, KRT18, and ALDH1A1) in the UKB population with available proximity extension assay (PEA) proteomic data. They compare subjects from different liver fibrosis severity stages, stratified via non-invasive liver fibrosis surrogates. A: Fibrosis-4 (FIB4) and B: AST-to-platelet ratio index (APRI). A Wilcoxon rank sum test was used to compare subgroups. Significance levels are indicated as follows: ns: not significant, **p* <0.05, ***p* <0.01, ****p* <0.001, *****p* <0.0001. *ADAMTSL2: ADAMTS-like protein 2; ALDH1A1: aldehyde dehydrogenase 1A1; IGFBP7: insulin-like growth factor-binding protein 7; ITGBL1: integrin beta-like protein 1; KRT18: keratin-18.*

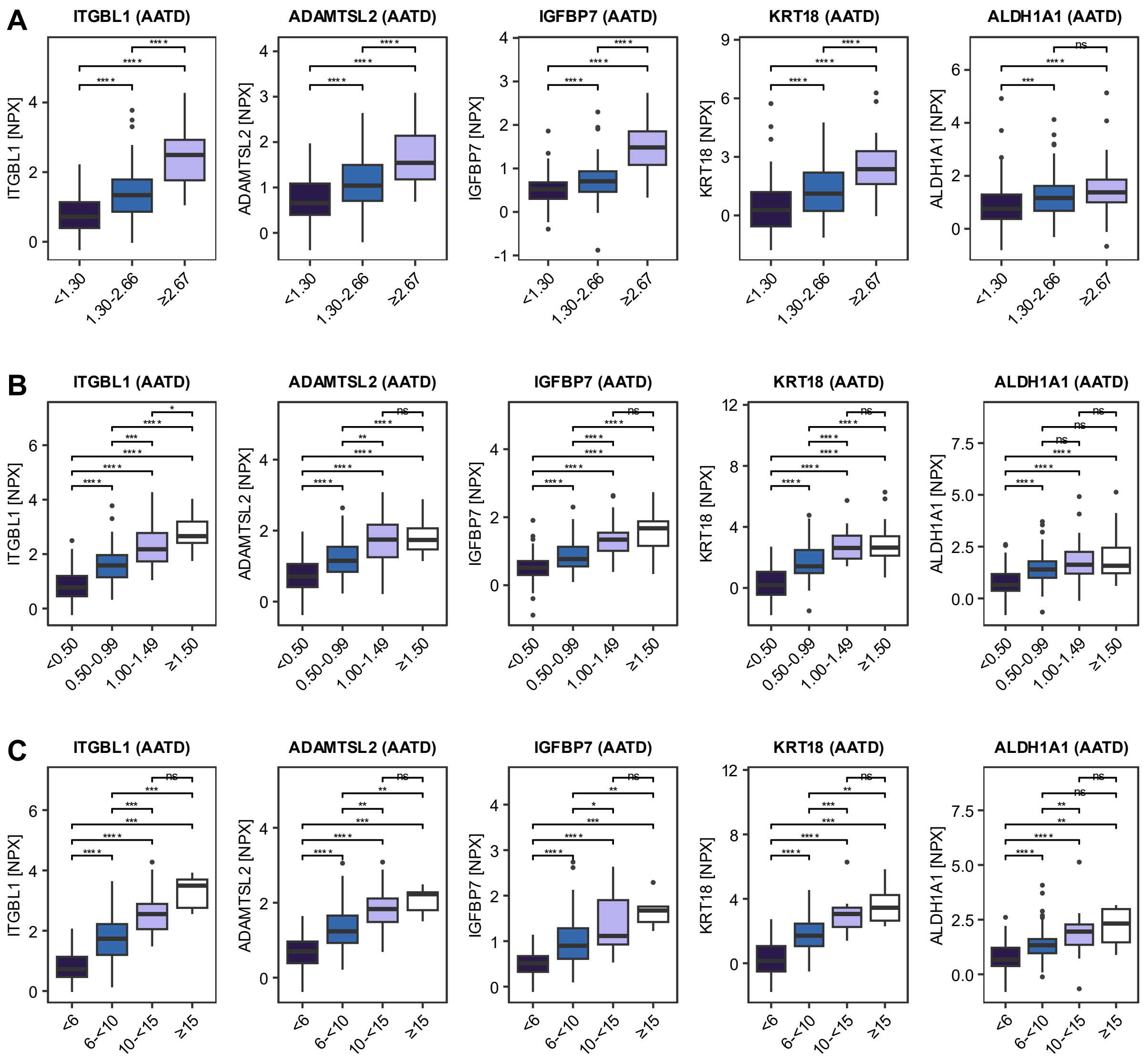

**Fig. S14: Ability of selected biomarkers to discriminate between liver fibrosis stages in a cohort of severe alpha1-antitrypsin deficiency (AATD) subjects.** Box plots display relative expression levels (NPX values) of five protein markers (ITGBL1, ADAMTSL2, IGFBP7, KRT18, and ALDH1A1) in the entire AATD cohort. They compare patients with different liver fibrosis severity stages, stratified via non-invasive liver fibrosis surrogates. A: Fibrosis-4 (FIB4), B: AST-to-platelet ratio index (APRI) and C: LiverRisk score. A Wilcoxon rank sum test was used to compare individual subgroups. Significance levels are indicated as follows: ns: not significant, **p* <0.05, ***p* <0.01, ****p* <0.001, *****p* <0.0001. *ADAMTSL2: ADAMTS-like protein 2; ALDH1A1: aldehyde dehydrogenase 1A1; IGFBP7: insulin-like growth factor-binding protein 7; ITGBL1: integrin beta-like protein 1; KRT18: keratin-18.*

### Supplementary Tables

**Table S1. List of ICD-10 and OPCS-4 codes.** Below are the ICD-10 and OPCS-4 codes used in this study. Codes marked with * are ascites codes that were only considered in combination with at least one chronic liver disease ICD-10 code (ICD-10: K70-K77). This assignment was used because ascites can have non-hepatic causes. ICD-10: Z944 was used to identify patients who received a liver transplant prior to baseline assessment and were excluded from the study. *ICD-10: International Classification of Diseases version 10, Tenth Revision; OPCS-4: operation/procedure codes version 4.*

| List of ICD-10 codes used | Description |
|---|---|
| K70.3 | Alcoholic cirrhosis of liver |
| K72.1 | Chronic hepatic failure |
| K74.6 | Other and unspecified cirrhosis of liver |
| K76.6 | Portal hypertension |
| K76.7 | Hepatorenal syndrome |
| I85.0; I859; I98.2; I98.3 | Oesophageal varices |
| I86.4 | Gastric varices |
| C22.0 | Hepatocellular carcinoma |
| Z94.4 | Liver transplant status |

| List of OPCS-4 codes used | Description |
|---|---|
| J06.1 | Transjugular intrahepatic insertion of stent into portal vein |
| J06.2 | Transjugular intrahepatic insertion of stent graft into portal vein |
| G10.4 | Local ligation of varices of oesophagus |
| G10.8 | Other specified open operations on varices of oesophagus |
| G10.9 | Unspecified open operations on varices of oesophagus |
| G14.4 | Fibreoptic endoscopic injection sclerotherapy to varices of oesophagus |
| G17.4 | Endoscopic injection sclerotherapy to varices of oesophagus using rigid oesophagoscope |
| G43.7 | Fibreoptic endoscopic rubber band ligation of upper gastrointestinal tract varices |
| T46.1* | Paracentesis abdominis for ascites |
| T46.2* | Drainage of ascites not elsewhere specified |

**Table S2. Demographics and clinical parameters of obese subjects (BMI>30 kg/m²) with available proteomic data from the UK Biobank cohort.** Cohorts of subjects who did vs. did not develop major adverse liver outcomes during the follow-up (MALO/non-MALO) are shown. Data are expressed as median (25$^{th}$-75$^{th}$ percentile) for continuous variables and n (%) for categorical variables. The p-values are obtained via ANOVA (* without covariates, ** covariates age, sex, and BMI were added). Parameters with p <1.00E-100 are highlighted in bold. *ALT: alanine aminotransferase; ALP: alkaline phosphatase; AST: aspartate aminotransferase; GGT gamma-glutamyltransferase; HbA1C: haemoglobin A1c.*

| Characteristics | **Non-MALOs**, N=12,737 | **MALOs**, N=228 | p-value |
|---|---|---|---|
| Age | 59 (51, 64) | 61 (56, 65) | 8.75E-07* |
| Sex (Female) | 6,746 (53%) | 71 (31%) | 5.97E-11* |
| BMI (kg/m$^2$) | 32.8 (31.2-35.5) | 33.4 (31.6-36.8) | 8.55E-05* |
| Type-2 Diabetes | 893 (7.0%) | 48 (21%) | 1.42E-16** |
| Blood glucose (mmol/L) | 5.04 (4.67-5.54) | 5.34 (4.81-6.58) | 7.61E-17** |
| HbA1C (mmol/mol) | 37 (34-40) | 39 (35-47) | 1.98E-08** |
| Total protein (g/L) | 72.4 (69.9-75.2) | 72.9 (70.1-76.6) | 5.57E-03** |
| Albumin (g/L) | 44.58 (42.86-46.28) | 43.31 (41.57-45.32) | 1.12E-15** |
| ALT (IU/L) | 24 (18-33) | 32 (21-50) | 2.15E-38** |
| **AST (IU/L)** | **25 (22-30)** | **36 (26-50)** | **1.86E-106**** |
| **GGT (IU/L)** | **34 (24-52)** | **88 (49-195)** | **8.66E-211**** |
| ALP (IU/L) | 86 (72-101) | 94 (79-120) | 1.84E-29** |
| Total bilirubin (μmol/L) | 7.7 (6.1-9.9) | 9.4 (6.9-12.8) | 5.94E-28** |
| Direct bilirubin (μmol/L) | 1.59 (1.29-2.06) | 2.13 (1.54-3.10) | 4.55E-73** |
| Platelets (10$^9$/L) | 252 (216-292) | 210 (160-254) | 2.50E-27** |

**Table S3. Demographics and clinical parameters of subjects with type-2 diabetes with available proteomic data from the UK Biobank cohort.** Cohorts of subjects who did vs. did not develop major adverse liver outcomes during the follow-up (MALO/non-MALO) are shown. Data are expressed as median ($25^{th}$-$75^{th}$ percentile) for continuous variables and n (%) for categorical variables. The p-values are obtained via ANOVA (*without covariates, **covariates age, sex, and BMI were added). *ALT: alanine aminotransferase; ALP: alkaline phosphatase; AST: aspartate aminotransferase; GGT: gamma-glutamyltransferase; HbA1C: haemoglobin A1c.*

| **Characteristics** | **Non-MALOs**, N = 1,535 | **MALOs**, N = 73 | **p-value** |
|---|---|---|---|
| Age | 62 (57-66) | 63 (61-67) | 2.21E-03* |
| Sex (Female) | 568 (37%) | 24 (33%) | 0.48* |
| BMI (kg/m$^2$) | 31.3 (27.5-35.2) | 32.1 (28.7-34.9) | 0.35* |
| Blood glucose (mmol/L) | 6.5 (5.3-8.8) | 7.0 (5.6-9.5) | 0.17** |
| HbA1C (mmol/mol) | 50 (43-59) | 51 (43-60) | 0.91** |
| Total protein (g/L) | 72.4 (69.5-75.4) | 73.8 (69.7-77.0) | 3.28E-02** |
| Albumin (g/L) | 44.7 (42.7-46.7) | 43.3 (40.1-45.5) | 1.18E-06** |
| ALT (IU/L) | 24 (18-34) | 28 (18-41) | 1.19E-04** |
| AST (IU/L) | 25 (21-31) | 34 (26-43) | 1.10E-14** |
| GGT (IU/L) | 35 (24-54) | 75 (45-186) | 6.74E-26** |
| ALP (IU/L) | 86 (71-103) | 97 (81-128) | 1.53E-05** |
| Total bilirubin (µmol/L) | 7.8 (6.1-10.1) | 8.7 (7.0-13.2) | 2.09E-08** |
| Direct bilirubin (µmol/L) | 1.74 (1.38-2.33) | 2.17 (1.53-3.26) | 2.49E-10** |
| Platelets ($10^9$/L) | 242 (205-292) | 210 (157-252) | 1.08E-07** |

**Table S4. Differential abundance analysis (Bayesian linear regression) comparing subjects from the UK Biobank cohort with available proximity extension assay (PEA) data with/without future major adverse liver outcomes (MALO).** A log2 fold-change >0 indicates proteins elevated in subjects with vs. without future MALO. 1772 proteins differed between both groups (FDR <0.05, 1330 elevated, 442 diminished in MALO). Data are sorted according to FDR. Tissue-specificities are indicated with a 0/1-coding (1: specificity for a given tissue). *logFC: log fold change; p: p-value; FDR: false discovery rate.*

**Table S5. Differential abundance analysis (Bayesian linear regression) comparing obese (BMI ≥30 kg/$m^2$) subjects from the UK Biobank with available proximity extension assay (PEA) data with/without future major adverse liver outcomes (MALO).** A log2 fold-change >0 indicates proteins elevated in subjects with vs. without future MALO. 1704 proteins differed between both groups (FDR <0.05, 1150 elevated, 554 diminished in MALO). Data are sorted according to FDR. Tissue-specificities are indicated with a 0/1-coding (1: specificity for a given tissue). *logFC: log fold change; p: p-value; FDR: false discovery rate.*

**Table S6. Differential abundance analysis (Bayesian linear regression) comparing Type-2 diabetic (DM2) subjects from the UK Biobank with available proximity extension assay (PEA) data with/without future major adverse liver outcomes (MALO).** A log2 fold-change >0 indicates proteins elevated in subjects with vs. without future MALO. 815 proteins differed between both groups (FDR <0.05, 745 elevated, 70 diminished in MALO). Data are sorted according to FDR. Tissue-specificities are indicated with a 0/1-coding (1: specificity for a given tissue). *logFC: log fold change; p: p-value; FDR: false discovery rate.*

**Table S7. Differential abundance analysis (Bayesian linear regression) comparing AATD subjects with vs. without significant liver fibrosis based on non-invasive liver stiffness measurement (LSM ≥7.0 kPa vs. <7.0 kPa).** A log2 fold-change >0 indicates proteins elevated in subjects with LSM ≥7.0 kPa vs. <7.0 kPa. 1266 proteins differed between both groups (FDR <0.05, 792 elevated, 470 diminished in subjects with LSM ≥7.0 kPa). Data are sorted according to FDR. Tissue-specificities are indicated with a 0/1-coding (1: specificity for a given tissue). *AATD: alpha1-antitrypsin deficiency; logFC: log fold change; p: p-value; FDR: false discovery rate.*

**Table S8. Differential abundance analysis (Bayesian linear regression) comparing AATD subjects with vs. without significant liver fibrosis based on non-invasive liver stiffness measurement (LSM 7.0-<15kPa vs. <7.0 kPa).** A log2 fold-change > 0 indicates proteins elevated in subjects with LSM of 7.0-<15kPa vs. <7.0 kPa. Subjects with possible clinically significant portal hypertension (LSM≥15) were excluded. 548 proteins differed between both groups (FDR <0.05, 442 elevated, 106 diminished in subjects with LSM of 7.0-<15kPa). Data are sorted according to FDR. Tissue-specificities are indicated with a 0/1-coding (1: specificity for a given tissue). *AATD: alpha1-antitrypsin deficiency; logFC: log fold change; p: p-value; FDR: false discovery rate.*

**Table S9: Demographic and routine parameters of people living with HIV (PLHIV).** Categorisation of subjects was based on non-invasive liver stiffness measurement (LSM), values are expressed in kPa. Data are expressed as median ($25^{th}$-$75^{th}$ percentile) for continuous variables and n (%) for categorical variables. The p-values are obtained via ANOVA (*without covariates, **with covariates age, sex, and BMI). *ALT: alanine aminotransferase; ALP: alkaline phosphatase; AST: aspartate aminotransferase; GGT: gamma-glutamyltransferase.*

| Characteristics | N | PLHIV cohort, Fibrosis grade based on LSM (kPa) | | p-value |
|---|---|---|---|---|
| | | **F0-F1** (<7.0) N = 874 | **≥F2** (≥7.0) N = 86 | |
| Age (years) | 960 | 52 (43-59) | 54 (41-62) | 0.541* |
| Sex (Female) | 960 | 120 (14%) | 8 (9.3%) | 0.250* |
| BMI (kg/m$^2$) | 960 | 24.9 (22.5-27.5) | 27.6 (24.3-30.9) | 8.54E-09* |
| Type-2 Diabetes | 960 | 38 (4.3%) | 13 (15%) | 1.49E-05** |
| ALT (IU/L) | 939 | 25 (20-33) | 31 (21-43) | 1.80E-09** |
| AST (IU/L) | 227 | 26 (21-31) | 28 (22-36) | 0.009** |
| GGT (IU/L) | 386 | 28 (18-49) | 33 (24-68) | 0.013** |
| ALP (IU/L) | 413 | 79 (66-94) | 82 (66-99) | 0.315** |
| Total bilirubin (µmol/L) | 768 | 7.0 (5.0-10.0) | 7.0 (6.0-10.0) | 0.028** |
| Platelets (10$^9$/L) | 957 | 216 (186-256) | 224 (183-275) | 0.126** |

**Table S10. Ability of selected biomarkers to predict MALOs in the UK Biobank (UKB) cohort.** The table displays areas under receiving operating curves (AUROCs) for the 20 proteins consistently associated with a significant liver disease in all studied cohorts (UKB, AATD and PLHIV) for the prediction of future major adverse liver outcomes (MALO) (logistic regression, age and sex added as covariates). AUROCs were calculated for all UKB participants with available proximity extension assay (PEA) data, as well as the obese (BMI ≥30 kg/m²) and diabetic (DM2) subgroups. *AATD: alpha1-antitrypsin deficiency; ACY1: aminoacylase 1; ADAMTSL2: ADAMTS-like protein 2; ADGRG1: adhesion G protein-coupled receptor G1; ALDH1A1: aldehyde dehydrogenase 1A1; ALT: alanine aminotransferase; ANGPT2: angiopoietin 2; AST: aspartate aminotransferase; CD80: CD80 molecule; CDH2: cadherin 2; CDH15: cadherin 15; CLSTN2: calsyntenin 2; DSC2: desmocollin 2; ENG: endoglin; ENPP2: ectonucleotide pyrophosphatase/phosphodiesterase 2; GGT: gamma-glutamyltransferase; IGF1: insulin-like growth factor 1; IGFBP7: insulin-like growth factor-binding protein 7; IGSF9: Immunoglobulin superfamily member 9; ITGBL1: integrin beta-like protein 1; KRT18: keratin-18; MENT: C1orf56 (chromosome 1 open reading frame 56); NFASC: neurofascin; PLHIV: people living with HIV; TFPI2: tissue factor pathway inhibitor 2; THBS2: thrombospondin-2.*

| | AUROCs | | | Differences in AUROCs | |
|---|---|---|---|---|---|
| | Overall PEA | Obese | DM2 | Overall PEA vs Obese | Overall PEA vs DM2 |
| **ADAMTSL2** | 0.809 | 0.830 | 0.817 | 0.021 | 0.008 |
| **IGFBP7** | 0.806 | 0.835 | 0.855 | 0.030 | 0.049 |
| **ITGBL1** | 0.804 | 0.837 | 0.867 | 0.033 | 0.064 |
| **THBS2** | 0.801 | 0.830 | 0.838 | 0.029 | 0.037 |
| **GGT (baseline, clinical chemistry)** | 0.801 | 0.819 | 0.800 | 0.018 | -0.001 |
| **NFASC** | 0.794 | 0.808 | 0.803 | 0.014 | 0.009 |
| **CD80** | 0.790 | 0.803 | 0.750 | 0.014 | -0.040 |
| **ADGRG1** | 0.782 | 0.792 | 0.775 | 0.010 | -0.007 |
| **KRT18** | 0.780 | 0.788 | 0.715 | 0.008 | -0.065 |
| **CDH2** | 0.773 | 0.785 | 0.764 | 0.012 | -0.009 |
| **ACY1** | 0.761 | 0.761 | 0.688 | 0.000 | -0.073 |
| **IGSF9** | 0.759 | 0.755 | 0.723 | -0.004 | -0.035 |
| **CLSTN2** | 0.756 | 0.796 | 0.810 | 0.040 | 0.054 |
| **AST (baseline, clinical chemistry)** | 0.754 | 0.769 | 0.728 | 0.015 | -0.026 |
| **ENPP2** | 0.751 | 0.781 | 0.762 | 0.030 | 0.010 |
| **ANGPT2** | 0.744 | 0.742 | 0.681 | -0.002 | -0.063 |
| **IGF1 (baseline, clinical chemistry)** | 0.743 | 0.762 | 0.740 | 0.020 | -0.002 |
| **ENG** | 0.729 | 0.767 | 0.739 | 0.038 | 0.010 |
| **MENT** | 0.729 | 0.753 | 0.729 | 0.024 | 0.000 |
| **DSC2** | 0.719 | 0.763 | 0.732 | 0.044 | 0.013 |
| **ALDH1A1** | 0.717 | 0.732 | 0.670 | 0.015 | -0.047 |
| **TFPI2** | 0.715 | 0.732 | 0.685 | 0.017 | -0.030 |
| **ALT (baseline, clinical chemistry)** | 0.711 | 0.706 | 0.613 | -0.005 | -0.098 |
| **CDH15** | 0.678 | 0.696 | 0.645 | 0.018 | -0.033 |
